# Six month repeat cognitive testing to identify people with MCI at greatest AD dementia risk

**DOI:** 10.64898/2025.12.30.25343228

**Authors:** Douglas Tommet, Nancy S. Foldi, Melissa Lamar, Chun Wang, Laura Rabin, Evan Grandoit, Seo-Eun Choi, Roos J. Jutten, Michael Lee, Connie Nakano, Laura E. Gibbons, Phoebe Scollard, Dan Mungas, Richard N. Jones, the Alzheimer’s Disease Neuroimaging Initiative, Paul K. Crane

**Author notes:** Correspondence to: Paul K. Crane, MD MPH, Box 359780, 325 Ninth Avenue, Seattle, WA 98106. Data used in preparation of this article were obtained from the Alzheimer’s Disease Neuroimaging Initiative (ADNI) database (adni.loni.usc.edu). As such, the investigators within the ADNI contributed to the design and implementation of ADNI and/or provided data but did not participate in analysis or writing of this report. A complete listing of ADNI investigators can be found at: http://adni.loni.usc.edu/wp-content/uploads/how_to_apply/ADNI_Acknowledgement_List.pdf.

## Abstract

**INTRODUCTION:** People with mild cognitive impairment (MCI) are candidates for early intervention, but not all progress to Alzheimer’s disease (AD) dementia. Identifying a subgroup at highest risk may improve treatment targeting.

**METHODS:** We analyzed data from participants with MCI enrolled in the Alzheimer’s Disease Neuroimaging Initiative (ADNI). Cognitive domains included memory, executive functioning, language, and visuospatial abilities. We evaluated baseline performance and 6-month change scores, using proportional hazards models to estimate associations with time to conversion to AD dementia.

**RESULTS:** The strength of association varied by domain, but in general both baseline performance and 6-month change were associated with conversion. The strongest effects observed for memory and language. Observed associations were largely independent of established risk biomarkers, including APOE genotype, structural MRI measures, and CSF biomarkers.

**DISCUSSION:** 6-month change scores on cognitive tests may help identify a high-risk subgroup of persons with MCI likely to progress to AD dementia.

**RESEARCH IN CONTEXT:**

1. Systematic review. The authors reviewed the literature using traditional (e.g. PubMed) sources. There is a modest literature on change scores in the context of the AD clinical spectrum, but few investigations have evaluated whether short-term changes may be able to identify a high-risk subgroup of people with MCI. The authors have published a systematic review of this literature (Jutten et al. 2020) and appropriately refer to relevant citations here.
2. Interpretation: Our findings suggest that short-term changes in cognition may be useful as part of a strategy to identify subsets of people with MCI who are at highest risk of conversion. Findings were clearest for memory and language. Domain-specific changes appeared to be independent from other biomarkers used to identify people at highest risk. Domain-specific changes did not appear to be better than changes in global cognition as measured by the MMSE or the CDR-sum of boxes.
3. Future directions: Short-term changes in cognition may be useful to help identify a subgroup of people with MCI at highest risk of conversion to AD dementia. Future work could consider time frames shorter than the 6-month data we had available, better characterizing changes with more than 2 time points, or developing strategies that combine changes in cognition with other biomarkers to identify a subgroup of people with MCI to target for treatment.

## 1.0 BACKGROUND

Recent approvals of anti-amyloid medications have ushered in a new era of management decisions for people with mild cognitive impairment (MCI) who could possibly be treated with these medications. Concerns about toxicity and resource allocation prompt some urgency in targeting treatment to those most likely to benefit.^1–4^

Several approaches have been evaluated to try to improve the identification of those at highest risk of converting from MCI to Alzheimer’s disease (AD) dementia, including considering data from cognitive testing, genetics, fluid and imaging biomarkers, and neuropsychiatric characteristics.^5–13^ Many of these efforts have focused on data measured at a single time point. Yet neurodegenerative processes leading to dementia evolve over time. Longitudinal data may provide information on evolution of processes over time, which may facilitate differentiation of people on a trajectory that will lead to dementia from people on other trajectories. Evaluation of short-term longitudinal data may prove to be an important strategy to identify people to treat and people not to treat. We sought to determine whether change in cognition over a six-month period might be a useful adjunct to other strategies to identify those with MCI who are at highest risk for subsequent progression to AD dementia. We focused on people who were known to be amyloid positive, analogous to people who would otherwise be eligible to be treated with anti-amyloid medications, because anti-amyloid medications are only indicated for people with demonstrated amyloid.

A second line of investigation was whether cognitive change associations with dementia risk were independent of findings from other types of biomarkers. Cognitive assessment is less expensive and is better tolerated than other modalities proposed to help identify those at greatest risk of conversion from MCI to AD dementia. Understanding whether a potential cognitive change biomarker is independent of other biomarkers is therefore important in developing strategies that balance tradeoffs involved with delaying therapy onset and the complexity of needing multiple assessment time points to identify those to treat.

Finally, we were curious as to whether the domain-specific approach was valuable for identifying people at greatest risk of conversion from MCI to AD dementia relative to measures of global cognition and clinical staging. To address this, we also analyzed global scores from the Mini-Mental State Examination (MMSE),^14^ the Alzheimer’s Disease Assessment Scale-Cognitive subscale (ADAS-Cog),^15^ and the Clinical Dementia Rating (CDR) scale’s “sum of boxes”.^16^

The goals of our analyses were first to determine whether short-term changes in memory, executive functioning, language, visuospatial, and global cognitive scores were associated with risk of subsequent conversion from MCI to AD dementia, and additionally among people with MCI and known to be amyloid positive using data from the AD Neuroimaging Initiative (ADNI). A second goal was to determine whether any associations with changes in cognitive domains were independent of or complementary to other fluid and imaging biomarkers. And a third goal was to determine whether short-term changes in global cognition and clinical staging were associated with risk of conversion from MCI to AD dementia.

## 2.0 METHODS

### 2.01 Overview

Data used in the preparation of this article were obtained from the Alzheimer’s Disease Neuroimaging Initiative (ADNI) database (adni.loni.usc.edu), a large multi-site observational study of older adults followed closely to identify conversions from MCI to AD dementia. The ADNI was launched in 2003 as a public-private partnership, led by Principal Investigator Michael W. Weiner, MD. The primary goal of ADNI has been to test whether serial magnetic resonance imaging (MRI), positron emission tomography (PET), other biological markers, and clinical and neuropsychological assessment can be combined to measure the progression of MCI and early AD. For up-to-date information, see www.adni-info.org.

ADNI administered an extensive battery of cognitive tests that we have previously processed to determine composite scores for four cognitive domains: memory, executive functioning, language, and visuospatial.^17–19^ We considered people from any stage of ADNI who joined the study with MCI, who had all four cognitive domain scores at baseline and at 6 months, and who had not developed AD dementia at the 6-month visit. We used both baseline performance and change from enrollment to 6 months in our models. We analyzed AD conversion outcomes over more than 4 years of follow-up, on average. We used Cox proportional hazards models for time to AD dementia conversion, with the 6-month visit set as the baseline. We added fluid and imaging-based biomarkers to our cognitive domain score models to determine whether signals associated with 6-month changes in cognitive domain scores were providing complementary or independent information compared with the other types of biomarkers. We evaluated three global cognitive measures using the same analytic framework. We performed all analyses in the overall sample and restricted to the subset of participants determined to be amyloid positive within 12 months of enrollment by either CSF fluid biomarker analyses or by amyloid PET scan.

### 2.02 Participants

We included individuals who enrolled in the first or second cycle of ADNI or in the ADNI Grand Opportunities (GO) enrollment wave with an initial diagnosis of MCI (total = 903; ADNI1 n=397, ADNI2 n=343, ADNIGO n=129). We excluded people with missing cognitive data (n=53), and those who either missed the 6-month visit (n=59) or converted to AD at the 6-month visit (n=46), resulting in an analytic sample of n=764 individuals with MCI (ADNI1 n=358, ADNI2 n=291, ADNIGO n=115). This group was followed for 3,199 person-years (mean 4.2 years per person).

We also performed analyses on the subset of this group of people with MCI determined to be amyloid positive based on CSF measures, amyloid PET scans, or both. Amyloid data were available within 1 year of the first study visit for 583 people with MCI (76% of those with MCI). Of these 583 people with MCI who met our eligibility criteria, 408 were found to be amyloid positive (70% of those with known amyloid status).

### 2.03 Diversity, Equity, and Inclusion (DEI)

ADNI had no exclusion criteria for any group based on ethnicity or race. ADNI enrollment was characterized by over-representation of people with European ancestry. Current ADNI funding focuses specifically on enhancing diversity in new enrollees. The present analyses are of data from the earlier parts of the ADNI study.

### 2.04 Amyloid Positivity

There were 196 people with MCI who were amyloid positive based on CSF assessment only (Elecsys Aβ42 < 1098 pg/mL^20^); 34 who were amyloid positive based on PET findings including [11C]PiB (global cortical standardized uptake value ratio [SUVR] ≥ 1.517), [18F]florbetapir (global cortical SUVR ≥ 1.1018), or [18 F]florbetaben (global cortical SUVR ≥ 1.1119), and 178 who were amyloid positive based on both CSF and PET. There were 76 people with MCI who had discrepant amyloid status between CSF and PET; all were included as amyloid-positive individuals here. Of these, 57 were amyloid positive based on CSF but not PET, and 19 were amyloid positive based on PET but not CSF. These data are summarized in **Supplemental Figure S1**.

There were 175 people with MCI included in the initial analyses who were found to be amyloid negative using the same cutoffs. Another 126 people (all from ADNI1) were excluded from these analyses because their amyloid status was not determined at all, and 55 people (all from ADNI1) were excluded because their amyloid status was determined >12 months after their baseline visit.

### 2.05 Cognitive domain scores

We used memory, executive functioning, language, and visuospatial composite scores, as described previously.^17–19^ Briefly, cognitive assessment items and tasks were assigned to cognitive domains by an expert panel of neuropsychologists and a behavioral neurologist, and we used confirmatory factor analysis models to compute scores for each domain. These scores have been uploaded and are available at the Laboratory on Neuro Imaging (LONI) website (https://adni.loni.usc.edu/). We considered scores from the baseline and 6-month visits for each domain as our primary exposure of interest, and determined change over that 6 months by subtracting the baseline score from the 6-month score.^21^

We categorized change scores into five groups for each domain: improved substantially, improved a little, minimal change (reference category), worsened a little, and worsened substantially. We categorized these groups to avoid assuming linear relationships between changes and risk of conversion, and to facilitate separate comparisons of improvement and of decline. We derived categorizations in two ways. First, we considered the distribution of change scores and tried to identify reasonable thresholds from the distribution of the data points. Second, we used k-means clustering, where the algorithm identifies thresholds that simultaneously maximize within group similarity (i.e. change scores closest to each other within each group) and between group differences (i.e. change scores furthest from each other between groups). Overall results were similar with both methods. Because k-means clustering better balanced the number of individuals in each group, we used groups defined on this basis for our primary analyses.

We performed similar analyses of scores from the enrollment visit, and we used k-means clustering for those categories as well. We entered both enrollment visit performance category and change category into our models.

### 2.06 Global cognitive scores and clinical status

We used MMSE, ADAS-Cog, and CDR-sum of boxes total scores for analyses. We used k-means clustering for the global cognitive scores to form groups to facilitate comparisons. For the MMSE and the CDR-sum of boxes, because the categories with the greatest improvements were small (<10 people), we combined that category together with the group with the 2^nd^ greatest improvement; the MMSE and CDR-sum of boxes analyses thus had 4 groups rather than 5. The CDR global score is derived from the individual box scores that make up the CDR Sum of Boxes. Because a diagnosis of dementia corresponds to the CDR global score changing from 0.5 to 1.0, there is some circularity in examining changes in the Sum of Boxes. We report these results for comparison, but interpret them with this limitation in mind.

### 2.07 Fluid and imaging biomarkers

We considered several other variables to determine their impact on associations between groups defined by 6-month changes in cognitive domain scores and risk of conversion to AD dementia. In particular, we used *APOE* genotype (coded as ≥1 *APOE* ε4 allele vs. 0 ε4 alleles), MRI findings from Freesurfer analyses available from LONI, and presence of the AD signature defined on the basis of cerebrospinal fluid (CSF) biomarkers as defined by De Meyer et al.^22^ in the subset of individuals for whom CSF biomarker data were available. We considered multiple relevant MRI-derived biomarkers including one global biomarker (cortical gray matter), memory-related biomarkers (hippocampal volume, entorhinal cortical thickness), language-related biomarkers (temporal lobe cortical thickness, medial temporal lobe cortical thickness, lateral temporal lobe cortical thickness), executive functioning-related biomarkers (frontal lobe cortical thickness, cingulate cortical thickness), visuospatial-related biomarkers (parietal lobe cortical thickness, occipital lobe cortical thickness) and a biomarker not thought to be related to any of these domains (sensory / motor cortical thickness) as a negative control.

### 2.08 Criteria to identify conversion to AD

ADNI’s approach to diagnosing conversion to AD is an operationalization of the NINCDS-ADRA (McKhann) criteria for probable or possible Alzheimer’s Disease,^23^ and is described in the ADNI protocol ^24^ Diagnoses at each visit were made by site clinicians using standardized clinical interviews (including the CDR), neuropsychological testing (ADAS-Cog, MMSE, and the full battery), functional and behavioral measures (FAQ, GDS, NPI-Q), adverse events, and laboratory data. An experienced physician integrated all available information to determine the diagnosis (normal, MCI, AD, or other) and to classify AD as Probable or Possible according to operationalized NINCDS-ADRDA criteria. Probable AD required objective memory impairment, decline in at least one additional cognitive domain, evidence of progressive deterioration, and no alternative primary cause. Possible AD was assigned when AD was considered the primary etiology, but a coexisting condition could also contribute to cognitive impairment. A Central Review Committee verified eligibility and all diagnostic conversions to ensure uniform application across sites.

### 2.09 Data analysis

We used Cox proportional hazards models to determine associations between enrollment visit performance groups and change score groups and risk of subsequent conversion from MCI to AD dementia. The Cox models used the 6 month visit as the starting point for time to capture differences in risk of conversion to AD dementia. All models in all analyses included terms for age, years of education, and female sex. We performed each analysis with two samples – the entire sample of 764 people, and again in the 408 people known to be amyloid positive. **Supplemental Figure S2** summarizes the modeling strategy.

Our initial models considered each domain separately (memory, executive functioning, language, and visuospatial). We included terms for groups defined by the enrollment visit score (with the middle group as the reference category) and for 6-month change (with the middle “minimal change” group excluded as the reference category). We used five categories for both baseline and change for each domain, except for visuospatial where we considered 4 categories of change due to poorer spread of the change scores compared to other domains.

We also considered the effects of biomarker covariates. Biomarker covariates were entered into the analysis models one at a time, with the goal of evaluating whether the effect estimated for baseline cognition or change category was altered by the inclusion of the covariate. We considered *APOE* genotype, various imaging-based parameters selected a priori, and the AD CSF biomarker signature. These analyses included the same categories for enrollment cognition performance and for changes between enrollment and the 6-month visit as for the primary models, except for the CSF biomarker signature analyses, where we combined categories due to the smaller sample size with available data. Additionally, to determine whether the baseline and change effects were specific for a cognitive domain, we performed additional analyses including baseline groups and 6-month change groups for all the cognitive domains in the same model (the above models included just one domain at a time). Finally, we considered global cognition scores and clinical status, with analysis models constructed similarly to the individual cognitive domains. We included categories for baseline scores along with categories for changes. The middle category was excluded for the baseline scores. We selected 4 categories to characterize changes in MMSE and CDR-sum of boxes. To maximize parallelism with the cognitive domain score analyses, we used the 3^rd^ category as the reference category. For the cognitive domain score analyses, the value of 0 (indicating no change at all) was included in that 3^rd^ category, so the “minimal change” label applied to all of those analyses. For the MMSE, the 3^rd^ category was −1 to −3, meaning the people in that middle reference category all declined from 1-3 points. Similarly, for the CDR-sum of boxes, the middle group included people whose sum of boxes totals were higher by 0.5 to 1.5 points.

We performed model checking of the proportional hazards assumption and investigated influential observations for our primary models in the entire dataset and in the amyloid positive subset. Analyses were conducted using R, version 4.2.2 (R Foundation for Statistical Computing, Vienna, Austria).^25^

### 2.10 Standard Protocol Approvals and Patient Consents

All ADNI participants signed IRB-approved consent forms. The University of Washington IRB approval is STUDY00008205.

## 3.0 RESULTS

The analyses included 764 participants with MCI across all amyloid status groups (amyloid status unknown, confirmed amyloid negative, and confirmed amyloid positive), of whom 408 were confirmed to be amyloid positive by either CSF biomarker levels or amyloid PET scans within 12 months of enrollment.

### 3.1 Categories Defined Based On Changes Between Enrollment And 6 Months

We categorized participants based on changes between the enrollment visit and the 6-month visit for each domain score. The thresholds used and the numbers in each category are shown in **Table 1**. We used similar approaches to categorize participants based on performance at the enrollment visit (**Supplemental Table S1**).

**Table 1.**
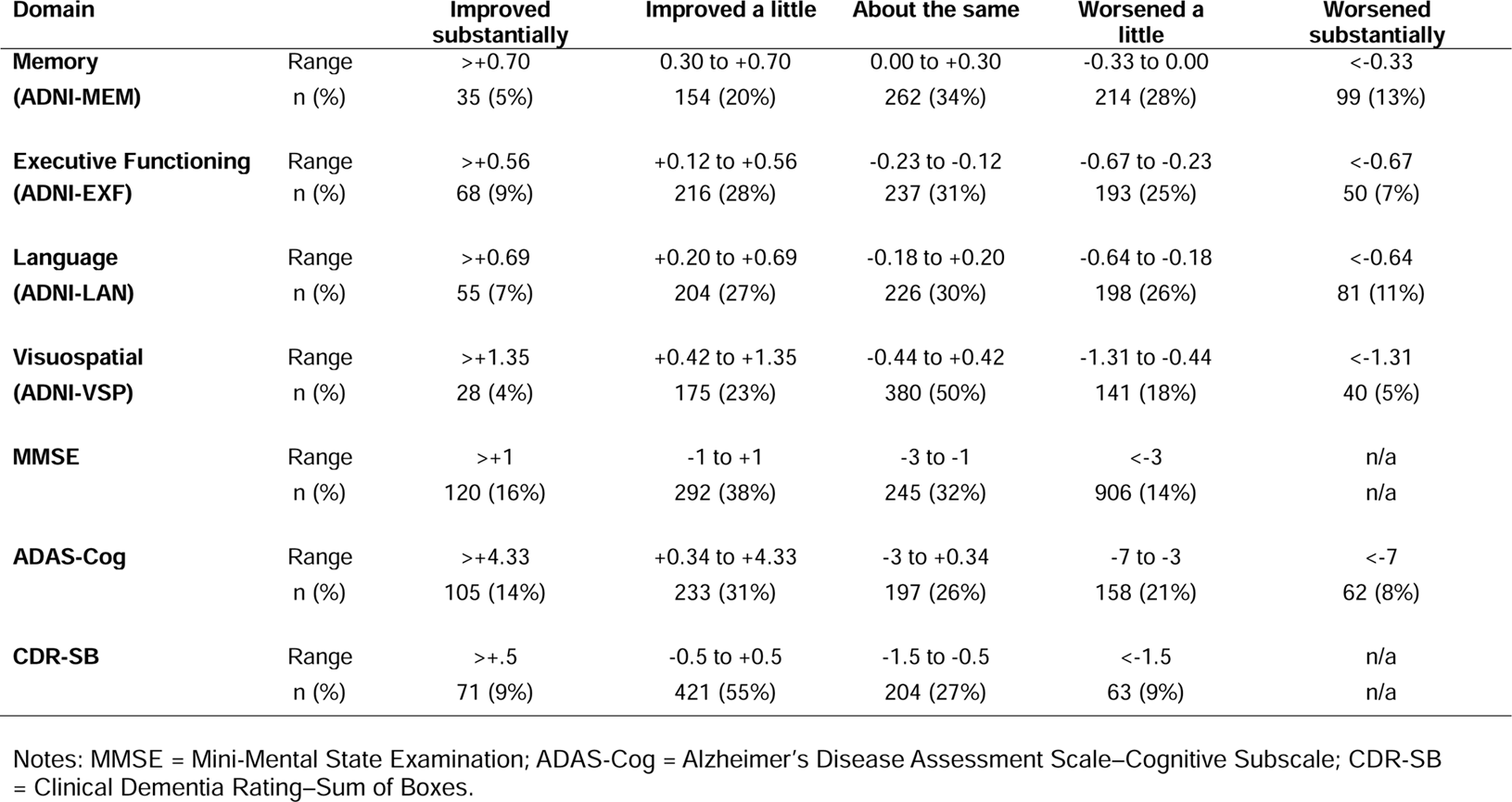
Performance categories for changes between enrollment and 6-month visit and numbers in each category, all participants (n=764)

### 3.2 Demographic Characteristics

In **Table 2** we provide an overview of the sociodemographics for the entire sample (n = 764), and by subgroups based on categories of change for the language domain. The overall sample had a mean age at enrollment of 72.7 years. This mean ranged from 71.8 to 74.1 across groups; the differences across groups had a p value of 0.4. In all, 41% of the overall sample were women, the mean years of education was 20.0 years, 94% self-reported white race, 9% were left-handed, and 50% had at least one *APOE* ε4 allele. None of these characteristics differed significantly across groups defined based on changes in language scores from enrollment to the 6-month visit. The proportion of people with at least one *APOE* ε4 allele was somewhat lower in the group whose language scores improved substantially (18/55 = 33%) compared to the overall sample (385/764 = 50%), though the chi squared *p* value for all five groups was not significant (*p*=0.09).

**Table 2.**
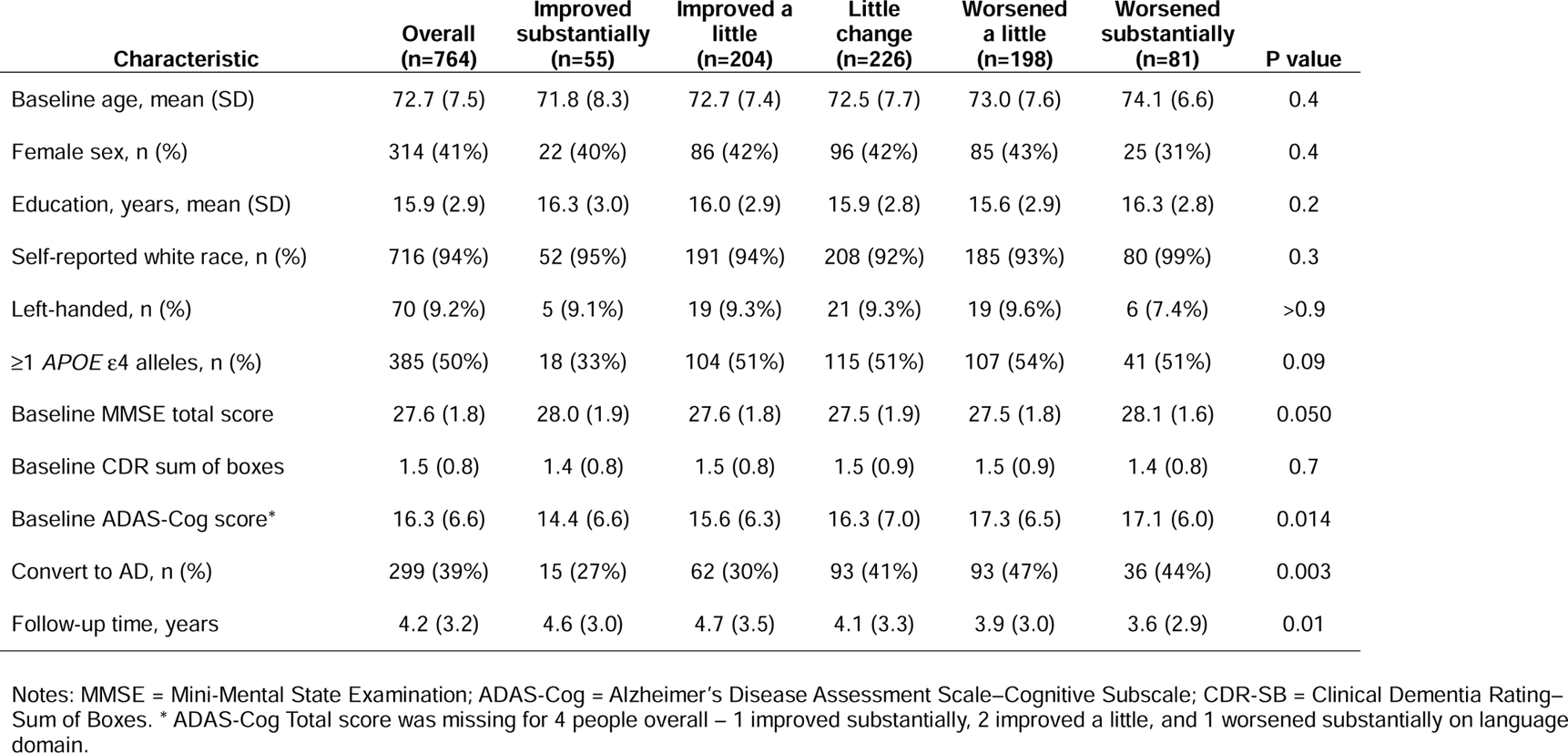
Demographic and clinical characteristics stratified by change in the language domain, all participants (n=764)

Baseline MMSE scores varied significantly (*p*=0.05) across groups, but differences were small and mean scores were lowest (worse) in the three middle groups, with mean scores the highest (best) in the “improved substantially” and “worsened substantially” groups. Overall cognitive and functional impairment, as measured by the Clinical Dementia Rating Scale Sum of Boxes, was very similar across the groups; however, the baseline cognition as measured by the ADAS-Cog total score differed across these groups, with lower (better) average scores for groups with improvement or minimal change compared to groups with worsening language scores.

Over a mean of 4.2 years of follow-up for this sample of 764 people with MCI, 299 (39%) converted to AD dementia. Across groups defined on the basis of changes in language scores between enrollment and the 6-month visit, the proportion who convert to AD dementia varied substantially, ranging from 27% in the “improved substantially” group to 47% in the “worsened a little” and 44% in the “worsened substantially” group; the unadjusted p value based on the chi squared statistic for this comparison was 0.003. These findings are dissected further with the Cox models reported below.

Demographic characteristics for the amyloid positive subset are shown in **Supplemental Table S1**. Relationships appeared similar in this subset of the data, except for *APOE* genotype. *APOE* ε4 alleles were more common among those who were amyloid positive (249/408 or 61% had ≥1 ε4 allele), and the proportion with at least one ε4 allele was lowest among those whose language scores improved substantially (10/31 = 32%) and highest among those whose language scores worsened substantially (34/49 = 69%), with a p value of 0.013. Among people who were positive for amyloid, MMSE scores did not differ across groups defined by 6-month changes in language score (p=0.3). ADAS-Cog total scores, CDR sum of boxes, and conversion to AD dementia findings across groups defined by 6-month changes in language score were similar to the pattern of findings observed in the whole sample.

### 3.3 Overlap In Changes Across Domains

We assessed the overlap across change scores for each cognitive domain. Correlations in changes from the enrollment visit to the 6 month visit for the four domain scores ranged from 0.06 to 0.17 (**Supplemental Table S3**). We used weighted Cohen’s kappa statistics to summarize agreement beyond chance in categories defined based on changes between enrollment and 6 months defined for each cognitive domain as shown in **Table 1**. The six weighted kappa statistics for groups defined based on changes between enrollment and 6 months for the four cognitive domains ranged from 0.03 to 0.09, where values <0.20 are considered “slight agreement” (see **Supplemental Table S4**). We used similar approaches to categorize people based on changes between enrollment and the 6-month visit for the MMSE, ADAS-Cog, and CDR sum of boxes, as shown in **Table 1**. Weighted kappa statistics describing agreement beyond chance for the three global scores ranged from 0.05 to 0.08. Weighted kappa statistics for eleven of the twelve comparisons of the four domain scores and the three global measures ranged from 0.01 to 0.10. The only weighted kappa statistic that was >0.20 was the one from the agreement beyond chance for changes in the ADAS-Cog and changes in memory, which was 0.22 (**Supplemental Table S4**).

### 3.4 Cognitive Domain Associations With Risk of Subsequent Conversion From MCI To AD Dementia

Results from Cox time-to-event models for each domain considered separately are shown in **Table 3**. For memory, for the entire cohort of people with MCI (left column of memory findings in Table 3 in gray, marked “All”), performance in memory from the enrollment visit was an important predictor of conversion from MCI at the 6-month visit to subsequent AD dementia. The group that had substantially improved memory scores between the enrollment visit and the 6-month visit also had a lower hazard ratio for conversion from MCI at the 6-month visit to subsequent AD dementia. For the subset known to be amyloid positive (right column for memory findings in Table 2 in white, marked “Amyloid +”), findings were similar, though lower hazards for conversion from MCI at the 6 month visit to subsequent AD dementia were seen both for the group with substantial improvement as well as the group with who improved a little.

**Table 3.** Hazard ratios for conversion to Alzheimer’s disease among those with Mild Cognitive Impairment in ADNI given baseline and change category membership in four domains of cognitive performance (domains considered separately)

|  | Memory |  | Executive Functioning |  | Language |  | Visuospatial |  |
| --- | --- | --- | --- | --- | --- | --- | --- | --- |
|  | All<br>(n = 764) | Amyloid +<br>(n=408) | All<br>(n = 764) | Amyloid +<br>(n=408) | All<br>(n = 764) | Amyloid +<br>(n=408) | All<br>(n = 764) | Amyloid +<br>(n=408) |
| **6-month change categories** |  |  |  |  |  |  |  |  |
| Improved substantially HR, 95% CI | **0.24**<br>**(0.09, 0.66)** | **0.16**<br>**(0.04, 0.65)** | **0.49**<br>**(0.31, 0.79)** | **0.57**<br>**(0.33, 0.98)** | **0.50**<br>**(0.29, 0.87)** | **0.45**<br>**(0.23, 0.89)** | 0.74<br>(0.39, 1.41) | 0.84<br>(0.39, 1.80) |
| Improved a little HR, 95% CI | 0.74<br>(0.53, 1.03) | **0.56**<br>**(0.37, 0.86)** | **0.74**<br>**(0.55, 1.00)** | 0.80<br>(0.54, 1.19) | **0.58**<br>**(0.42, 0.81)** | **0.53**<br>**(0.35, 0.80)** | **0.58**<br>**(0.42, 0.80)** | 0.67<br>(0.44, 1.02) |
| Minimal change (reference) | 1 (reference) | 1 (reference) | 1 (reference) | 1 (reference) | 1 (reference) | 1 (reference) | 1 (reference) | 1 (reference) |
| Worsened a little HR, 95% CI | 1.10<br>(0.83, 1.45) | 0.97<br>(0.68, 1.39) | 1.17<br>(0.87, 1.58) | **1.56**<br>**(1.07, 2.29)** | 1.33<br>(0.99, 1.77) | 1.05<br>(0.73, 1.52) | 1.30<br>(0.95, 1.76) | 1.28<br>(0.88, 1.88) |
| Worsened substantially HR, 95% CI | 1.39<br>(0.96, 2.00) | 1.24<br>(0.77, 2.00) | 1.39<br>(0.86, 2.26) | 1.76<br>(0.94, 3/29) | **2.28**<br>**(1.51, 3.46)** | **2.02**<br>**(1.19, 3.43)** | **1.65**<br>**(1.03, 2.63)** | **1.92**<br>**(1.09, 3.37)** |
| **Baseline value categories** |  |  |  |  |  |  |  |  |
| High HR, 95% CI | **0.17**<br>**(0.10, 0.29)** | **0.20**<br>**(0.10, 0.39)** | **0.37**<br>**(0.24, 0.57)** | **0.37**<br>**(0.21, 0.66)** | **0.26**<br>**(0.16, 0.41)** | **0.28**<br>**(0.16, 0.51)** | **0.49**<br>**(0.37, 0.66)** | **0.57**<br>**(0.40, 0.83)** |
| Medium high HR, 95% CI | **0.49**<br>**(0.32, 0.73)** | **0.49**<br>**(0.29, 0.82)** | 0.69<br>(0.47, 1.01) | 0.71<br>(0.44, 1.15) | 0.71<br>(0.49, 1.02) | 0.68<br>(0.44, 1.05) | n/a | n/a |
| Medium (reference) | 1 (reference) | 1 (reference) | 1 (reference) | 1 (reference) | 1 (reference) | 1 (reference) | 1 (reference) | 1 (reference) |
| Medium low HR, 95% CI | **1.82**<br>**(1.30, 2.54)** | **1.66**<br>**(1.10, 2.52)** | 1.31<br>(0.93, 1.85) | 1.21<br>(0.78, 1.88) | 1.00<br>(0.70, 1.43) | 0.95<br>(0.60, 1.50) | 1.20<br>(0.84, 1.73) | 1.11<br>(0.68, 1.79) |
| Low HR, 95% CI | **3.60**<br>**(2.61, 4.97)** | **2.85**<br>**(2.61, 4.97)** | **3.04**<br>**(2.17, 4.26)** | **2.64**<br>**(1.71, 4.07)** | **2.21**<br>**(1.56, 3.12)** | **2.22**<br>**(1.44, 3.42)** | 1.33<br>(0.93, 1.90) | 1.43<br>(0.92, 2.23) |
| **Demographic characteristics** |  |  |  |  |  |  |  |  |
| Age HR per year, 95% CI | 1.02<br>(1.00, 1.03) | 1.01<br>(0.99, 1.03) | 1.01<br>(0.99, 1.03) | 1.00<br>(0.98, 1.02) | 1.01<br>(1.00, 1.03) | 1.00<br>(0.98, 1.02) | **1.03**<br>**(1.02, 1.05)** | 1.01<br>(0.99, 1.04) |
| Female sex HR, 95% CI | **1.44**<br>**(1.13, 1.82)** | **1.78**<br>**(1.30, 2.44)** | 1.12<br>(0.88, 1.42) | **1.38**<br>**(1.02, 1.86)** | 1.22<br>(0.96, 1.55) | 1.28<br>(0.95, 1.73) | 1.05<br>(0.82, 1.33) | 1.23<br>(0.91, 1.66) |
| Education HR per year, 95% CI | 1.03<br>(0.99, 1.07) | 1.03<br>(0.97, 1.08) | 1.04<br>(1.00, 1.08) | 1.00<br>(0.94, 1.05) | **1.05**<br>**(1.01, 1.09)** | 1.02<br>(0.97, 1.08) | 1.01<br>(0.97, 1.05) | 0.97<br>(0.92, 1.03) |

Note: Table entries are hazard ratios (HR) with 95% confidence limits derived from Cox proportional hazards models.

For executive functioning, performance at the enrollment visit was an important predictor of conversion from MCI at the 6-month visit to subsequent AD dementia, especially for the extreme categories, for both the entire cohort and the subset known to be amyloid positive. For the entire cohort, the groups with substantial improvement and with a little improvement between the enrollment visit and the 6-month visit had lower hazard ratios for conversion from MCI at the 6-month visit to subsequent AD dementia. For the amyloid positive subset, point estimates were similar for the group with a little improvement between enrollment and the 6-month visit, and the group with substantial improvement had lower hazard ratios for conversion from MCI at the 6-month visit to subsequent AD dementia. In the amyloid positive subset, the group with a little worsening between the enrollment visit and the 6-month visit had a higher hazard ratio for conversion from MCI at the 6-month visit to subsequent AD.

For language, performance at the enrollment visit was an important predictor of conversion from MCI at the 6-month visit to subsequent AD dementia, especially for the extreme categories, for both the entire cohort and the subset known to be amyloid positive. For both the entire cohort and the amyloid positive subset, both the improved a little and improved substantially groups had lower hazard ratios for conversion from MCI at the 6-month visit to subsequent AD dementia, and the substantially worsened group had higher hazard ratios for conversion from MCI at the 6-month visit to subsequent AD dementia.

For visuospatial, we pooled together all high scores from baseline in a single category. People in this category had a lower risk of conversion to AD dementia in the entire cohort and the subset known to be amyloid positive. The group who improved a little between enrollment and the 6-month visit had a lower hazard ratio for conversion from MCI at the 6-month visit to subsequent AD dementia for the entire cohort, and the substantially worsened group had a higher hazard ratio for conversion from MCI at the 6-month visit to subsequent AD dementia for both the entire cohort and for the subset known to be amyloid positive.

Results from single Cox models that included categories defined based on enrollment visit performance and on changes between enrollment visit and the 6-month visit for all four domains are shown in **Table 4**. Performance for memory at the enrollment visit was particularly important as a predictor of conversion from MCI at the 6-month visit to subsequent AD dementia both in the entire cohort (all 4 groups had hazard ratios that were significantly different from the null, with the middle group as the reference category) and among those known to be amyloid positive (3 of the 4 groups had hazard ratios that were significantly different from the null compared to the middle group as the reference category). The group with the lowest baseline executive functioning scores also had a higher hazard ratio for conversion to AD dementia compared to the middle group for the entire cohort and for those known to be amyloid positive. People whose memory scores improved substantially between the enrollment and 6-month visits had a lower hazard ratio for conversion from MCI at the 6-month visit to subsequent AD dementia in the entire cohort and among those known to be amyloid positive. In addition, people whose memory scores improved a little between the enrollment visit and the 6-month visit also had a lower hazard ratio for conversion to AD dementia compared to the group with minimal change among those known to be amyloid positive. People whose language scores improved a little between the enrollment visit and the 6-month visit also had lower hazard ratios for conversion from MCI at the 6-month visit to subsequent AD dementia compared to the group with minimal change for the entire cohort and for those known to be amyloid positive.

**Table 4.** Hazard ratios for conversion to Alzheimer’s disease among those with Mild Cognitive Impairment in ADNI given baseline and change category membership in four domains of cognitive performance (domains considered simultaneously)

|  | Memory |  | Executive Functioning |  | Language |  | Visuospatial |  |
| --- | --- | --- | --- | --- | --- | --- | --- | --- |
|  | All<br>(n = 764) | Amyloid +<br>(n=408) | All<br>(n = 764) | Amyloid +<br>(n=408) | All<br>(n = 764) | Amyloid +<br>(n=408) | All<br>(n = 764) | Amyloid +<br>(n=408) |
| **month change categories** |  |  |  |  |  |  |  |  |
| Improved substantially HR, 95% CI | **0.29**<br>**(0.11, 0.82)** | **0.14**<br>**(0.03, 0.60)** | 0.64<br>(0.40, 1.04) | 0.84<br>(0.47, 1.50) | 0.74<br>(0.42, 1.30) | 0.72<br>(0.35, 1.48) | 1.37<br>(0.71, 2.64) | 1.80<br>(0.81, 3.99) |
| Improved a little HR, 95% CI | 0.72<br>(0.50, 1.04) | **0.47**<br>**(0.29, 0.77)** | 0.87<br>(0.64, 1.19) | 0.93<br>(0.60, 1.44) | **0.63**<br>**(0.45, 0.88)** | **0.56**<br>**(0.36, 0.89)** | 0.76<br>(0.54, 1.06) | 0.96<br>(0.60, 1.52) |
| Minimal change (reference) | 1 (reference) | 1 (reference) | 1 (reference) | 1 (reference) | 1 (reference) | 1 (reference) | 1 (reference) | 1 (reference) |
| Worsened a little HR, 95% CI | 1.03<br>(0.77, 1.37) | 0.83<br>(0.56, 1.24) | 1.05<br>(0.76, 1.44) | 1.50<br>(0.99, 2.26) | 1.02<br>(0.75, 1.38) | 0.88<br>(0.60, 1.29) | 0.90<br>(0.65, 1.24) | 0.89<br>(0.59, 1.34) |
| Worsened substantially HR, 95% CI | 1.29<br>(0.89, 1.88) | 1.16<br>(0.71, 1.89) | 1.32<br>(0.79, 2.19) | 1.58<br>(0.82, 3.07) | 1.23<br>(0.79, 1.91) | 0.88<br>(0.49, 1.56) | 0.85<br>(0.51, 1.42) | 1.34<br>(0.73, 2.46) |
| **baseline values** |  |  |  |  |  |  |  |  |
| High HR, 95% CI | **0.20**<br>**(0.11, 0.36)** | **0.22**<br>**(0.11, 0.47)** | 0.67<br>(0.42, 1.06) | 0.88<br>(0.47, 1.64) | 0.63<br>(0.39, 1.03) | 0.64<br>(0.34, 1.21) | 0.88<br>(0.63, 1.22) | 0.83<br>(0.54, 1.27) |
| Medium high HR, 95% CI | **0.55**<br>**(0.36, 0.84)** | 0.61<br>(0.35, 1.07) | 0.93<br>(0.63, 1.39) | 1.17<br>(0.71, 1.96) | 0.90<br>(0.61, 1.33) | 0.81<br>(0.50, 1.32) | n/a | n/a |
| Medium (reference) | 1 (reference) | 1 (reference) | 1 (reference) | 1 (reference) | 1 (reference) | 1 (reference) | 1 (reference) | 1 (reference) |
| Medium low HR, 95% CI | **1.65**<br>**(1.17, 2.32)** | **1.83**<br>**(1.15, 2.91)** | 1.15<br>(0.80, 1.64) | 1.16<br>(0.72, 1.86) | 0.76<br>(0.52, 1.11) | 0.65<br>(0.39, 1.07) | 0.99<br>(0.67, 1.47) | 0.63<br>(0.37, 1.09) |
| Low HR, 95% CI | **2.80**<br>**(1.95, 4.02)** | **3.12**<br>**(1.94, 5.02)** | **1.83**<br>**(1.25, 2.68)** | **1.87**<br>**(1.13, 3.07)** | 1.00<br>(0.67, 1.48) | 1.08<br>(0.64, 1.83) | 0.83<br>(0.57, 1.21) | 0.72<br>(0.44, 1.17) |
| **demographic characteristics** |  |  |  |  |  |  |  |  |
| Age HR per year, 95% CI | 1.01<br>(0.99, 1.02) | 1.01<br>(0.99, 1.04) |  |  |  |  |  |  |
| Female sex HR, 95% CI | **1.51**<br>**(1.18, 1.94)** | **1.78**<br>**(1.29, 2.46)** |  |  |  |  |  |  |
| Education HR per year, 95% CI | **1.06**<br>**(1.02, 1.11)** | 1.04<br>(0.98, 1.10) |  |  |  |  |  |  |

Note: Table entries are hazard ratios (HR) with 95% confidence limits derived from Cox proportional hazards models. This table summarizes the association of baseline and six-month change in cognitive domain scores with conversion to AD estimated in models that include all four domains in a single model. The effects of demographics are included under the Memory head but are not particular to memory.

There were no concerns about the proportional hazards assumption in the amyloid-positive subset (**Supplemental Results 1**). In the full cohort analyses, several variables showed improved model fit when a time-dependent (non-linear) term was added. Follow-up evaluations did not identify individual observations within those groups that were driving these findings (**Supplemental Results 2**).

### 3.4 Analyses Adding Other Biomarkers To The Cognitive Domain Association Models

We performed additional analyses to determine whether these associations were consistent when adding additional predictors of conversion to the Cox models, including *APOE* genotype, several MRI-derived biomarkers, and the presence of the CSF biomarker-derived AD signature. For the memory domain, the pattern of findings was not altered substantially with the addition of *APOE* genotype, cortical gray matter volume, hippocampal volume, entorhinal cortical thickness, temporal cortical thickness, medial temporal lobe cortical thickness, lateral temporal lobe cortical thickness, frontal lobe cortical thickness, cingulate cortical thickness, parietal lobe cortical thickness, occipital lobe cortical thickness, or sensory/motor cortical thickness (**Supplemental Tables S6-S9**). Similarly, for the executive functioning domain, the pattern of findings was not altered substantially with the addition of any of these covariates (**Supplemental Tables S10-S13**). For the language domain, the only covariates that had much of an impact on the pattern of findings was with language-related imaging findings, where the findings for groups with the lowest scores at enrollment were attenuated when each imaging finding was also included in the model (**Supplemental Tables S14 and S15**). The pattern of findings was not altered substantially with any of the other covariates (**Supplemental Tables S16 and S17**) For the visuospatial domain, the pattern of findings was not altered substantially with the addition of any of the covariates (**Supplemental Tables S18-S21**).

Smaller numbers of participants had CSF-derived biomarkers, which led us to collapse several of the groups together for the enrollment visit and for the change between enrollment and the 6-month visit. Findings with this smaller sample were similar to those from the whole sample, though with wider confidence intervals. Including the CSF-based biomarker signature had minimal impact on the pattern of findings for any of the domains (**Supplemental Tables S22-S25**).

### 3.5 Global Cognitive Scores And Clinical Status Associations With Risk Of Conversion From MCI To Ad Dementia

We performed additional analyses in which we considered three scores summarizing global cognition or clinical status. We used the same settings for categorizing groups based on results from the enrollment visit and for changes between the enrollment and 6-month visits, as shown in **Table 1**.

Results from Cox time-to-event models for the MMSE, ADAS-Cog, and CDR-sum of boxes are shown in **Table 5**. For each of these, groups defined based on findings at the enrollment visit were important predictors of risk for conversion from MCI at the 6-month visit to subsequent AD dementia, both for the entire cohort and for the amyloid positive subset. For each of these measures, changes in scores between the enrollment visit and the 6-month visit were also important predictors of risk of conversion from MCI at the 6-month visit and subsequent AD dementia.

**Table 5.** Cox model results for each global cognition score separately.

|  | MMSE<br>(4 categories) |  | ADAS-Cog |  | CDR-SB |  |
| --- | --- | --- | --- | --- | --- | --- |
|  | All<br>(n = 764) | Amyloid +<br>(n=408) | All<br>(n = 764) | Amyloid +<br>(n=408) | All<br>(n = 764) | Amyloid +<br>(n=408) |
| **6-month change categories** |  |  |  |  |  |  |
| Improved substantially HR, 95% CI | **0.22**<br>**(0.14, 0.33)** | **0.21**<br>**(0.13, 0.36)** | **0.53**<br>**(0.36, 078)** | **0.61**<br>**(0.37, 0.99)** | **0.27**<br>**(0.17, 0.43)** | **0.31**<br>**(0.18, 0.54)** |
| Improved a little HR, 95% CI | **0.67**<br>**(0.51, 0.88)** | **0.59**<br>**(0.42, 0.84)** | **0.60**<br>**(0.44, 0.84)** | 0.73<br>(0.48, 1.12) | **0.42**<br>**(0.32, 0.54)** | **0.46**<br>**(0.33, 0.65)** |
| Minimal change (reference) | 1 (reference) | 1 (reference) | 1 (reference) | 1 (reference) | 1 (reference) | 1 (reference) |
| Worsened a little HR, 95% CI | **1.44**<br>**(1.04, 1.99)** | 1.40<br>(0.93, 2.11) | **1.40**<br>**(1.91, 1.95)** | **1.62**<br>**(1.07, 2.46)** | **1.65**<br>**(1.13, 2.40)** | **2.71**<br>**(1.68, 4.37)** |
| Worsened substantially HR, 95% CI |  |  | **2.46**<br>**(1.62, 3.75)** | **3.15**<br>**(1.81, 5.49)** |  |  |
| **Baseline value categories** |  |  |  |  |  |  |
| High HR, 95% CI | **0.57**<br>**(0.35, 0.92)** | 0.59<br>(0.32, 1.09) | **3.96**<br>**(2.82, 5.55)** | **3.24**<br>**(2.11, 4.99)** | **2.84**<br>**(2.05, 3.94)** | **2.09**<br>**(1.41, 3.10)** |
| Medium high HR, 95% CI | 0.86<br>(0.58, 1.28) | 0.99<br>(0.60, 1.62) | **1.87**<br>**(1.31, 2.66)** | **1.73**<br>**(1.08, 2.75)** | 1.32<br>(0.92, 1.90) | 1.10<br>(0.69, 1.78) |
| Medium (reference) | 1 (reference) | 1 (reference) | 1 (reference) | 1 (reference) | 1 (reference) | 1 (reference) |
| Medium low HR, 95% CI | **3.06**<br>**(2.14, 4.36)** | **3.02**<br>**(1.90, 4.81)** | **0.52**<br>**(0.34, 0.79)** | **0.53**<br>**(0.31, 0.90)** | **0.67**<br>**(0.47, 0.94)** | **0.47**<br>**(0.31, 0.73)** |
| Low HR, 95% CI | **4.05**<br>**(2.72, 6.04)** | **3.71**<br>**(2.25, 6.12)** | **0.19**<br>**(0.11, 0.32)** | **0.24**<br>**(0.12, 0.49)** | **0.37**<br>**(0.25, 0.57)** | **0.25**<br>**(0.15, 0.43)** |
| **Demographic characteristics** |  |  |  |  |  |  |
| Age HR per year, 95% CI | 1.01<br>(1.00, 1.03) | 1.01<br>(0.99, 1.03) | 1.01<br>(0.99, 2.02) | 1.00<br>(0.98, 1.02) | **1.03**<br>**(1.01, 1.04)** | 1.02<br>(1.00, 1.04) |
| Female sex HR, 95% CI | 1.06<br>(0.84, 1.35) | 1.19<br>(0.88, 1.61) | 1.21<br>(0.95, 1.54) | 1.23<br>(0.90, 1.67) | 1.09<br>(0.86, 1.38) | 1.24<br>(0.92, 1.67) |
| Education HR per year, 95% CI | 1.01<br>(0.97, 1.06) | 1.00<br>(0.94, 1.05) | 1.04<br>(1.00, 1.08) | 0.99<br>(0.94, 1.04) | 0.98<br>(0.94, 1.02) | **0.93**<br>**(0.88, 0.98)** |

Note: Abbreviations: HR hazard ratio, CI Confidence interval; MMSE = Mini-Mental State Examination; ADAS-Cog = Alzheimer's Disease Assessment Scale–Cognitive Subscale; CDR-SB = Clinical Dementia Rating–Sum of Boxes.

## 4.0 DISCUSSION

We evaluated levels of cognition at study entry and short-term changes in four cognitive domains of memory, executive functioning, language, and visuospatial functioning. We did not find strong agreement in changes across domains. We found levels of cognition at study enrollment and changes between enrollment and 6 months were able to identify groups of people at differential risk of conversion from MCI to subsequent AD dementia. These relationships were especially notable for memory and language, not only in the analyses of each individual domain (**Table 2**) but also considering all domains together in a single analysis (**Table 3**). Findings were minimally impacted when including *APOE* genotype, MRI-based biomarkers, or the AD CSF biomarker signature. Findings were similar for the entire cohort and for the subset who were known to be amyloid positive. We also evaluated three tests of global cognition or clinical status. For each of these, we found that both baseline values and 6-month change were strongly associated with conversion risk (**Table 4**). All models included categories defined based on the baseline scores for each measure, such that any risk associated with groups defined by 6-month changes was beyond that implied by the enrollment visit score alone. In sum, the 6-month change scores appear to be able to identify groups of people at highest risk of dementia.

For each cognitive domain, participants who improved substantially over 6 months were at lower risk of conversion form MCI to AD compared to people with minimal change. For the language domain, participants with little improvement also appeared to have lower risk. With this similar pattern of findings across domains, we wondered whether these associations could be driven by a group of people with substantial improvement across all domains with a low conversion rate. However, the kappa statistic analyses showed little agreement beyond chance in categories defined by changes across domains. Furthermore, we included groups defined by changes in all four domains in a single model. Risk of conversion associated with substantial improvement in memory was independent, as the strength of association was very similar with groups defined by changes in other domains included. Risks for groups defined based on changes in executive functioning and language were somewhat attenuated in the combined model, though the association for the group with a little improvement in language was still statistically significant. Including *APOE* genotype or several MRI-based indicators did not substantially impact findings, suggesting change scores provide independent and complementary information on conversion risk. There was a smaller sample of people with CSF biomarker data. With this small sample size, the signal for improvements in memory appeared to be substantially attenuated by including the CSF biomarker-derived AD signature, and the signal for improvement in language appeared stronger.

The finding that changes in the memory and language domains provide independent information about risk of subsequent conversion is useful in providing context for findings regarding global cognition or clinical indicators. With global scores or with domain specific composite scores, two time points seemed valuable in stratifying risk of conversion from MCI to AD dementia. The domain-specific further analyses suggest language and memory aspects of these global indicators may be particularly relevant. Further research may be needed to further tease apart the global versus domain-specific nature of these relationships.

In a previous systematic review of the literature on change scores as potential indicators of subsequent risk for transition from MCI to AD, we did not find other research papers that had considered multiple domains as we have here.^26^ In general, the visuospatial domain did not appear as helpful as the other domains in identifying people at different risk for conversion. We previously showed the visuospatial domain is measured with much less measurement precision than the other cognitive domains in ADNI.^27^ In the context of a noisier measure such as visuospatial functioning, there may be signal we are not able to observe.

ADNI preferentially enrolled people with amnestic MCI with or without impairments in other domains. The strong signal for 6-month improvements in memory is intriguing. The findings for 6-month improvements in language were also quite interesting. While the primary role of AD on the memory domain is well documented, there is substantial heterogeneity in patterns of deficits across other domains. The group whose language scores improve substantially or the larger group of people whose scores improve a little bit both had lower risks of subsequent AD dementia conversion.

Our initial rationale was to consider ways to identify subsets of people with MCI at differential risk of subsequent AD dementia. These investigations are perhaps even more relevant in considering ways to identify individuals with MCI who may be most likely to benefit from anti-amyloid therapies that have considerable expense and risk of toxicity. Cognitive testing is noninvasive and relatively inexpensive especially compared to other biomarkers. Our findings suggest that samples could be enriched for people at higher risk for conversion by excluding those with substantial improvements in memory and those whose language scores over 6 months were substantially improved or a little improved. This strategy could be combined with approaches based on other biomarkers, as these factors appeared to be independent.

Highly relevant for the question of who with MCI might benefit from anti-amyloid treatments, we performed all analyses among people who were known to be amyloid positive. Patterns of findings were identical for this group. FDA-approved anti-amyloid treatments should only be offered to people who are, at minimum, amyloid positive. Not everyone with MCI who is amyloid positive progresses to AD dementia. Longitudinal cognitive data may provide important hints as to who may be most likely to progress to AD dementia without an intervention, and to identify those who would be unlikely to progress to AD dementia. Longitudinal cognitive data from amyloid positive people with MCI may assist in prioritizing treatment resources targeting potentially toxic anti-amyloid treatments to those individuals at highest risk of progression.

Our findings should be considered alongside strengths and limitations. While ADNI’s sample is large and drawn from dozens of research centers spread over the US and Canada, the sample represents a highly selected group of volunteers willing to consent to extensive periodic investigations with substantial participant burden. The ADNI sample on average is highly educated and relatively homogenous ethno-racially (similar to the clinical trial data to date on anti-amyloid therapies); however, the current cycle of ADNI – and the larger clinical trials field – is focused on enhancing the diversity of the sample, which will help with the generalizability of findings. ADNI’s cognitive battery is extensive but as mentioned above its measures of the visuospatial domain are characterized by more imprecision than seen in other domains; the lack of an association for 6-month changes in visuospatial scores may be due to relatively noisy visuospatial measurement in ADNI. While ADNI has massive sample sizes compared to many other studies of people with MCI, the sample with CSF data was too small for strong conclusions, and required us to combine groups to ensure adequate sample sizes in each of the groups analyzed. We considered several covariates, but as in any observational study there is the distinct possibility of unmeasured and residual confounding. Finally, the data we considered to determine changes in cognition were limited to between enrollment and 6-months follow-up. Whether the improved precision afforded by additional assessments would further enhance our ability to identify people at the highest risk of subsequent conversion from MCI to AD dementia is worthy of further study.

In conclusion, we found that 6-month improvements in memory and in language as well as in executive functioning were associated with lower risk of subsequent conversion to AD for people who enrolled with MCI in ADNI. Similarly, we found that 6-month improvement or worsening in global cognition scores were associated with differential risk of subsequent conversion to AD for people who enrolled with MCI in ADNI. Those wishing to enrich samples of people with MCI to ensure the enrolled group is at highest risk of conversion may wish to consider including 6-month changes in cognition as part of an enrichment strategy.

## Abbreviations

AD: Alzheimer’s disease
ADNI: AD Neuroimaging Initiative
CSF: Cerebral Spinal Fluid
DEI: Diversity, Equity, and Inclusion
ADNIGO: ADNI Grand Opportunity
MCI: Mild Cognitive Impairment

## Data Availability

Data used in the preparation of this article were obtained from the Alzheimer’s Disease Neuroimaging Initiative (ADNI) database (adni.loni.usc.edu), a large multi-site observational study of older adults followed closely to identify conversions from MCI to AD dementia.

http://adni.loni.usc.edu

## Acknowledgements

Acknowledgements

Data collection and sharing for this project was funded by the Alzheimer’s Disease Neuroimaging Initiative (ADNI) (National Institutes of Health Grant U01 AG024904) and DOD ADNI (Department of Defense award number W81XWH-12-2-0012). ADNI is funded by the National Institute on Aging, the National Institute of Biomedical Imaging and Bioengineering, and through generous contributions from the following: AbbVie, Alzheimer’s Association; Alzheimer’s Drug Discovery Foundation; Araclon Biotech; BioClinica, Inc.; Biogen; Bristol-Myers Squibb Company; CereSpir, Inc.; Cogstate; Eisai Inc.; Elan Pharmaceuticals, Inc.; Eli Lilly and Company; EuroImmun; F. Hoffmann-La Roche Ltd and its affiliated company Genentech, Inc.; Fujirebio; GE Healthcare; IXICO Ltd.;Janssen Alzheimer Immunotherapy Research & Development, LLC.; Johnson & Johnson Pharmaceutical Research & Development LLC.; Lumosity; Lundbeck; Merck & Co., Inc.;Meso Scale Diagnostics, LLC.; NeuroRx Research; Neurotrack Technologies; Novartis Pharmaceuticals Corporation; Pfizer Inc.; Piramal Imaging; Servier; Takeda Pharmaceutical Company; and Transition Therapeutics. The Canadian Institutes of Health Research is providing funds to support ADNI clinical sites in Canada. Private sector contributions are facilitated by the Foundation for the National Institutes of Health (www.fnih.org). The grantee organization is the Northern California Institute for Research and Education, and the study is coordinated by the Alzheimer’s Therapeutic Research Institute at the University of Southern California. ADNI data are disseminated by the Laboratory for Neuro Imaging at the University of Southern California.

## Conflicts

No declarations of interest.

## Funding sources

Analyses for this project were supported by NIH R01AG 029672 (P Crane, PI) and NIGMS SC3 GM122622 (N Foldi, PI) Data collection and sharing for this project was funded by the Alzheimer’s Disease Neuroimaging Initiative (ADNI) (National Institutes of Health Grant U01 AG024904) and DOD ADNI (Department of Defense award number W81XWH-12-2-0012). ADNI is funded by the National Institute on Aging, the National Institute of Biomedical Imaging and Bioengineering, and through generous contributions from the following: AbbVie, Alzheimer’s Association; Alzheimer’s Drug Discovery Foundation; Araclon Biotech; BioClinica, Inc.; Biogen; Bristol-Myers Squibb Company; CereSpir, Inc.; Cogstate; Eisai Inc.; Elan Pharmaceuticals, Inc.; Eli Lilly and Company; EuroImmun; F. Hoffmann-La Roche Ltd and its affiliated company Genentech, Inc.; Fujirebio; GE Healthcare; IXICO Ltd.;Janssen Alzheimer Immunotherapy Research & Development, LLC.; Johnson & Johnson Pharmaceutical Research & Development LLC.; Lumosity; Lundbeck; Merck & Co., Inc.;Meso Scale Diagnostics, LLC.; NeuroRx Research; Neurotrack Technologies; Novartis Pharmaceuticals Corporation; Pfizer Inc.; Piramal Imaging; Servier; Takeda Pharmaceutical Company; and Transition Therapeutics. The Canadian Institutes of Health Research is providing funds to support ADNI clinical sites in Canada. Private sector contributions are facilitated by the Foundation for the National Institutes of Health (www.fnih.org). The grantee organization is the Northern California Institute for Research and Education, and the study is coordinated by the Alzheimer’s Therapeutic Research Institute at the University of Southern California. ADNI data are disseminated by the Laboratory for Neuro Imaging at the University of Southern California.

## Notes

### Competing Interest Statement

The authors have declared no competing interest.

