## supplemental materials for "Six month repeat cognitive testing to identify people with MCI at greatest AD dementia risk"

### Supplemental Figure S1. Summary of CSF and imaging amyloid positive status

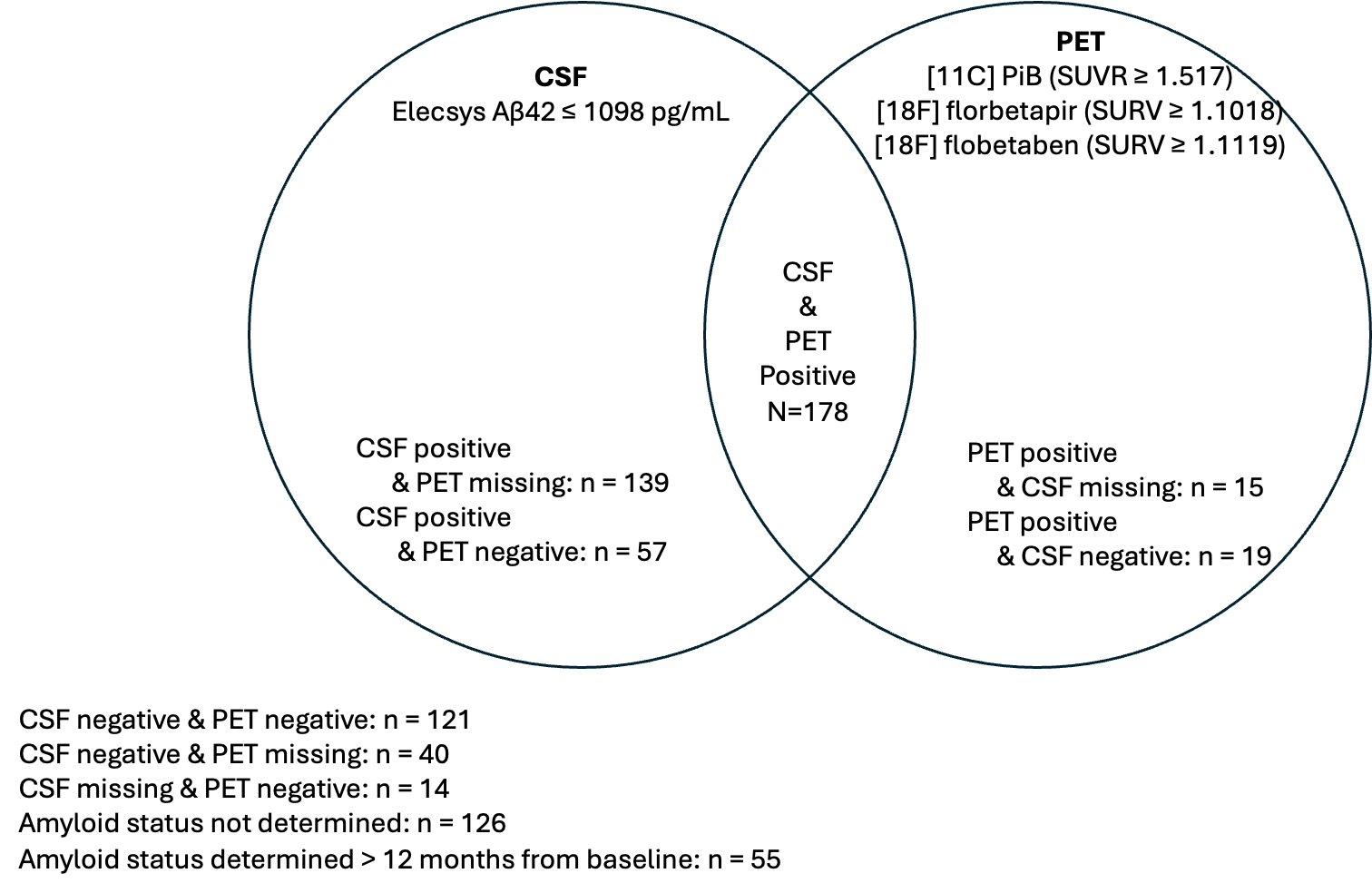

### Supplemental Figure S2. Summary of modeling approach*

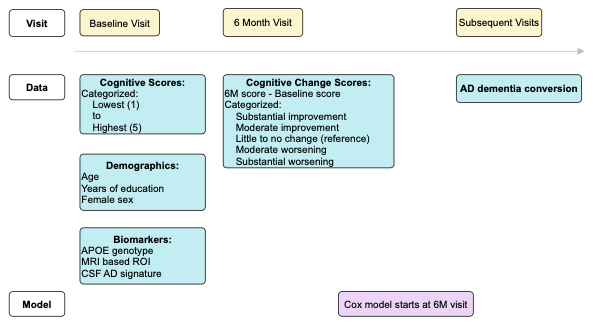

* Our primary variables of interest were domain scores at baseline and changes over 6 months (shown in blue). We categorized baseline scores and change scores. All models included terms for age, years of education, and female sex (shown in green). We then added an additional term to each model as shown in orange – *APOE* genotype, selected imaging-based parameters, and the CSF biomarker signature. We used this same approach for memory, executive functioning, language, and visuospatial. We performed all of these analyses with the entire sample of people with MCI at enrollment and the subset who were known to be amyloid positive within 12 months of enrollment. We considered time at risk beginning from the 6 month visit; our primary outcome was conversion to AD dementia (shown in gray).

### Supplemental Table S1. Thresholds for performance at the enrollment visit and numbers in each category.

| **Domain** |  | **Highest Scores** | | **2^nd^ Highest** | **Middle** | **4^th^ Highest** | **Lowest Scores** | |
| --- | --- | --- | --- | --- | --- | --- | --- | --- |
| **Memory** | Range | +0.73 to +2.28 | +0.30 to +0.72 | | -0.04 to +0.30 | -0.39 to -0.05 | | -1.53 to -0.39 |
|  | n (%) | 153 (20%) | 153 (20%) | | 153 (20%) | 153 (20%) | | 152 (20%) |
| **Executive Functioning** | Range | +0.73 to +2.41 | +0.28 to +0.72 | | -0.13 to +0.27 | -0.60 to -0.13 | | -2.54 to -0.61 |
|  | n (%) | 153 (20%) | 153 (20%) | | 153 (20%) | 153 (20 %) | | 152 (20%) |
| **Language** | Range | +0.89 to +2.69 | +0.42 to +0.89 | | +0.03 to +0.41 | -0.42 to +0.03 | | -2.88 to -0.42 |
|  | n (%) | 151 (20%) | 156 (20%) | | 152 (20%) | 153 (20%) | | 152 (20%) |
| **Visuospatial** | Range | +0.75 to +0.75 | -0.08 to -0.08 | | -0.74 to -0.24 | -2.77 to -0.86 | | n/a |
|  | n (%) | 298 (39%) | 221 (29%) | | 107 (14%) | 138 (18%) | | n/a |
| **MMSE** | Range | 30 | 29 | | 28 | 26-27 | | 23-24 |
|  | n (%) | 159 (20%) | 180 (23%) | | 139 (17%) | 199 (25%) | | 122 (15%) |
| **ADAS-Cog** | Range | 3 to 10 | 10 to 14 | | 14 to 18 | 18 to 22 | | 22 to 38 |
|  | n (%) | 156 (21%) | 146 (19%) | | 142 (19%) | 157 (21%) | | 159 (21%) |
| **CDR-SB** | Range | 0.5 | 1 | | 1.5 | 2 | | 2.5 to 5.5 |
|  | n (%) | 164 (21%) | 196 (26%) | | 168 (22%) | 114 (15%) | | 122 (16%) |

### Supplemental Table S2. Characteristics of the entire sample (gray columns) and the subset who were amyloid positive (white columns).

The data in the gray columns duplicates data in Table 2 in the main paper.

|  | **Overall** | | **Improved substantially** | | **Improved a little** | | **Little change** | | **Worsened a little** | | **Worsened substantially** | | **P value** | |
| --- | --- | --- | --- | --- | --- | --- | --- | --- | --- | --- | --- | --- | --- | --- |
| Characteristic | **All (n=764)** | **Amy + (n=408)** | **All (n=55)** | **Amy + (n=31)** | **All (n=204)** | **Amy + (n=98)** | **All (n=226)** | **Amy + (n=126)** | **All (n=198)** | **Amy + (n=104)** | **All (n=81)** | **Amy + (n=49)** | **All** | **Amy +** |
| Baseline age, mean (SD) | 72.7 (7.5) | 72.9 (7.2) | 71.8 (8.3) | 74.4 (7.4) | 72.7 (7.4) | 72.4 (7.0) | 72.5 (7.7) | 72.0 (7.6) | 73.0 (7.6) | 73.4 (7.2) | 74.1 (6.6) | 74.2 (6.1) | 0.4 | 0.3 |
| Female sex, n (%) | 314 (41%) | 167 (41%) | 22  (40%) | 10  (32%) | 86  (42%) | 45  (46%) | 96  (42%) | 51  (40%) | 85  (43%) | 47  (45%) | 25  (31%) | 14  (29%) | 0.4 | 0.2 |
| Education, years, mean (SD) | 15.9 (2.9) | 16.1 (2.8) | 16.3 (3.0) | 16.6 (3.0) | 16.0 (2.9) | 16.4 (2.7) | 15.9 (2.8) | 15.8 (2.8) | 15.6 (2.9) | 15.6 (2.8) | 16.3 (2.8) | 16.5 (2.9) | 0.2 | 0.12 |
| Self-reported white race, n (%) | 716 (94%) | 387 (95%) | 52  (95%) | 29  (94%) | 191 (94%) | 95  (97%) | 208 (92%) | 119 (94%) | 185 (93%) | 95  (91%) | 80  (99%) | 49 (100%) | 0.3 | 0.2 |
| Left handed, n (%) | 70 (9.2%) | 34 (8.3%) | 5  (9.1%) | 2  (6.5%) | 19 (9.3%) | 9  (9.2%) | 21 (9.3%) | 14  (11%) | 19 (9.6%) | 7  (6.7%) | 6  (7.4%) | 2  (4.1%) | >0.9 | 0.6 |
| ≥1 *APOE* ε4 alleles, n (%) | 385 (50%) | 249 (61%) | 18  (33%) | 10  (32%) | 104 (51%) | 61  (62%) | 115 (51%) | 78  (62%) | 107 (54%) | 66  (63%) | 41  (51%) | 34  (69%) | 0.09 | 0.013 |
| Baseline MMSE total score* | 27.6 (1.8) | 27.5 (1.8) | 28.0 (1.9) | 27.8 (1.8) | 27.6 (1.8) | 27.7 (1.9) | 27.5 (1.9) | 27.3 (1.9) | 27.5 (1.8) | 27.5 (1.8) | 28.1 (1.6) | 27.9 (1.7) | 0.050 | 0.3 |
| Baseline CDR sum of boxes* | 1.5  (0.8) | 1.5  (0.9) | 1.4  (0.8) | 1.3  (0.8) | 1.5  (0.8) | 1.6  (0.9) | 1.5  (0.9) | 1.5  (0.8) | 1.5  (0.9) | 1.6  (1.0) | 1.4  (0.8) | 1.3  (0.8) | 0.7 | 0.3 |
| Baseline ADAS-Cog score*# | 16.3 (6.6) | 17.3 (6.6) | 14.4 (6.6) | 15.6 (5.6) | 15.6 (6.3) | 15.8 (6.6) | 16.3 (7.0) | 18.0 (6.9) | 17.3 (6.5) | 17.8 (6.5) | 17.1 (6.0) | 18.2 (6.0) | 0.014 | 0.022 |
| Convert to AD, n (%) | 299 (39%) | 188 (46%) | 15  (27%) | 10  (32%) | 62  (30%) | 35  (36%) | 93  (41%) | 65  (52%) | 93  (47%) | 54  (52%) | 36  (44%) | 24  (49%) | 0.003 | 0.045 |
| Follow-up time, years | 4.2  (3.2) | 3.69 (2.71) | 4.6  (3.0) | 4.42 (2.82) | 4.7  (3.5) | 4.20 (2.81) | 4.1  (3.3) | 3.49 (2.67) | 3.9  (3.0) | 3.42 (2.48) | 3.6  (2.9) | 3.33 (2.92) | 0.01 | 0.026 |

### Supplemental Table S3. Thresholds between groups and numbers in each group for the entire sample

|  | **Memory** | **Executive functioning** | **Language** | **Visuospatial** | **MMSE** | **ADAS-Cog** | **CDR-SB** |
| --- | --- | --- | --- | --- | --- | --- | --- |
| **Baseline scores** |  |  |  |  |  |  |  |
| Highest |  |  |  |  |  |  |  |
| Range | +0.73 to +2.28 | +0.73 to +2.41 | +0.89 to +2.69 | +0.75 to +0.75 | 30 | 3 to 10 | 0.50 |
| N (%) | 153 (20%) | 153 (20%) | 151 (20%) | 298 (39%) | 159 (20%) | 156 (21%) | 164 (21%) |
| 2^nd^ highest |  |  |  |  |  |  |  |
| Range | +0.30 to +0.72 | +0.28 to +0.72 | +0.42 to +0.89 | -0.08 to -0.08 | 29 | 10 to 14 | 1.0 |
| N (%) | 153 (20%) | 153 (20%) | 156 (20%) | 221 (29%) | 180 (23%) | 146 (19%) | 196 (26%) |
| Middle |  |  |  |  |  |  |  |
| Range | -0.04 to +0.30 | -0.13 to +0.27 | +0.03 to +0.41 | -0.74 to -0.24 | 28 | 14 to 18 | 1.5 |
| N (%) | 153 (20%) | 153 (20%) | 152 (20%) | 107 (14%) | 139 (17%) | 142 (19%) | 168 (22%) |
| 4^th^ highest |  |  |  |  |  |  |  |
| Range | -0.39 to -0.05 | -0.60 to -0.13 | -0.42 to +0.03 | -2.77 to -0.86 | 26-27 | 18 to 22 | 2 |
| N (%) | 153 (20%) | 153 (20 %) | 153 (20%) | 138 (18%) | 199 (25%) | 157 (21%) | 114 (15%) |
| Lowest |  |  |  |  |  |  |  |
| Range | -1.53 to -0.39 | -2.54 to -0.61 | -2.88 to -0.42 |  | 23-24 | 22 to 38 | 2.5 to 5.5 |
| N (%) | 152 (20%) | 152 (20%) | 152 (20%) |  | 122 (15%) | 159 (21%) | 122 (16%) |
| **6-month change scores** |  |  |  |  |  |  |  |
| Improved substantially |  |  |  |  |  |  |  |
| Range | >+0.70 | >+0.56 | >+0.69 | >+1.35 | >+1 | >+4.33 | >+.5 |
| N (%) | 35 (5%) | 68 (9%) | 55 (7%) | 28 (4%) | 120 (16%) | 105 (14%) | 71 (9%) |
| Improved a little |  |  |  |  |  |  |  |
| Range | +0.30 to +0.70 | +0.12 to +0.56 | +0.20 to +0.69 | +0.42 to +1.35 | -1 to +1 | +0.34 to +4.33 | -0.5 to +0.5 |
| N (%) | 154 (20%) | 216 (28%) | 204 (27%) | 175 (23%) | 292 (38%) | 233 (31%) | 421 (55%) |
| Minimal change |  |  |  |  |  |  |  |
| Range | 0.00 to +0.30 | -0.23 to -0.12 | -0.18 to +0.20 | -0.44 to +0.42 | -3 to -1 | -3 to +0.34 | -1.5 to -0.5 |
| N (%) | 262 (34%) | 237 (31%) | 226 (30%) | 380 (50%) | 245 (32%) | 197 (26%) | 204 (27%) |
| Worsened a little |  |  |  |  |  |  |  |
| Range | -0.33 to 0.00 | -0.67 to -0.23 | -0.64 to -0.18 | -1.31 to -0.44 | <-3 | -7 to -3 | <-1.5 |
| N (%) | 214 (28%) | 193 (25%) | 198 (26%) | 141 (18%) | 906 (14%) | 158 (21%) | 63 (9%) |
| Worsened substantially |  |  |  |  |  |  |  |
| Range | <-0.33 | <-0.67 | <-0.64 | <-1.31 |  | <-7 |  |
| N (%) | 99 (13%) | 50 (7%) | 81 (11%) | 40 (5%) |  | 62 (8%) |  |

### Supplemental Table S4. Correlations of (continuous) change scores across domains

|  | **Change in Memory** | **Change in Executive Functioning** | **Change in Language** | **Change in Visuospatial** | **Change in MMSE** | **Change in ADAS-Cog** |
| --- | --- | --- | --- | --- | --- | --- |
| **Change in Memory** | 1 |  |  |  |  |  |
| **Change in Executive Functioning** | 0.14 | 1 |  |  |  |  |
| **Change in Language** | 0.15 | 0.17 | 1 |  |  |  |
| **Change in Visuospatial** | 0.10 | 0.12 | 0.06 | 1 |  |  |
| **Change in MMSE** | 0.21 | 0.10 | 0.12 | 0.11 | 1 |  |
| **Change in ADAS-Cog** | 0.40 | 0.15 | 0.15 | 0.10 | 0.18 | 1 |
| **Change in CDR-SB** | 0.12 | 0.07 | 0.13 | 0.05 | 0.14 | 0.12 |

### Supplemental Table S5. Weighted kappa statistics on agreement beyond chance for categories of change scores

|  | **Change in Memory** | **Change in Executive Functioning** | **Change in Language** | **Change in Visuospatial** | **Change in MMSE** | **Change in ADAS-Cog** |
| --- | --- | --- | --- | --- | --- | --- |
| **Change in Memory** | - |  |  |  |  |  |
| **Change in Executive Functioning** | 0.088 | - |  |  |  |  |
| **Change in Language** | 0.070 | 0.081 | - |  |  |  |
| **Change in Visuospatial** | 0.051 | 0.068 | 0.033 | - |  |  |
| **Change in MMSE** | 0.097 | 0.063 | 0.042 | 0.060 | - |  |
| **Change in ADAS-Cog** | 0.217 | 0.071 | 0.069 | 0.054 | 0.078 | - |
| **Change in CDR-SB** | 0.050 | 0.036 | 0.055 | 0.008 | 0.062 | 0.045 |

Weighted kappa coefficients using quadratic weights.

### Supplemental Results 1. Model checking for the amyloid positive subset.

We performed additional analyses to check the proportional hazards assumption. Each variable is added to the model with a non-linear term. The p value column in this table shows results of tests of whether that non-linear term is statistically different from the null. All of these p values were >0.05 for the amyloid positive group for the model with all four of the cognitive domains included, and provide little evidence that the proportional hazards assumption is violated for the listed covariates.

| **Covariate** | **Chi-square** | **df** | **p** |
| --- | --- | --- | --- |
| Baseline memory category (ADNI MEM) | 5.30 | 4 | 0.26 |
| Baseline executive functioning category (ADNI EFN) | 2.96 | 4 | 0.56 |
| Baseline language functioning category (ADNI LAN) | 5.26 | 4 | 0.26 |
| Baseline visuospatial functioning category (ADNI VSP) | 5.28 | 3 | 0.15 |
| Memory score (ADNI MEM) change score categories | 7.67 | 4 | 0.10 |
| Executive Functioning score (ADNI EFN) change score categories | 5.87 | 4 | 0.21 |
| Language functioning (ADNI LAN) change score categories | 1.89 | 4 | 0.76 |
| Visuospatial functioning (ADNI VSP) change score categories | 0.34 | 4 | 0.99 |
| Age | 2.44 | 1 | 0.12 |
| Gender | 3.12 | 1 | 0.08 |
| Educational attainment | 0.51 | 1 | 0.48 |
| Global test, all covariates | 41.80 | 34 | 0.17 |

We also evaluated Martingale residuals and found no particular outliers:

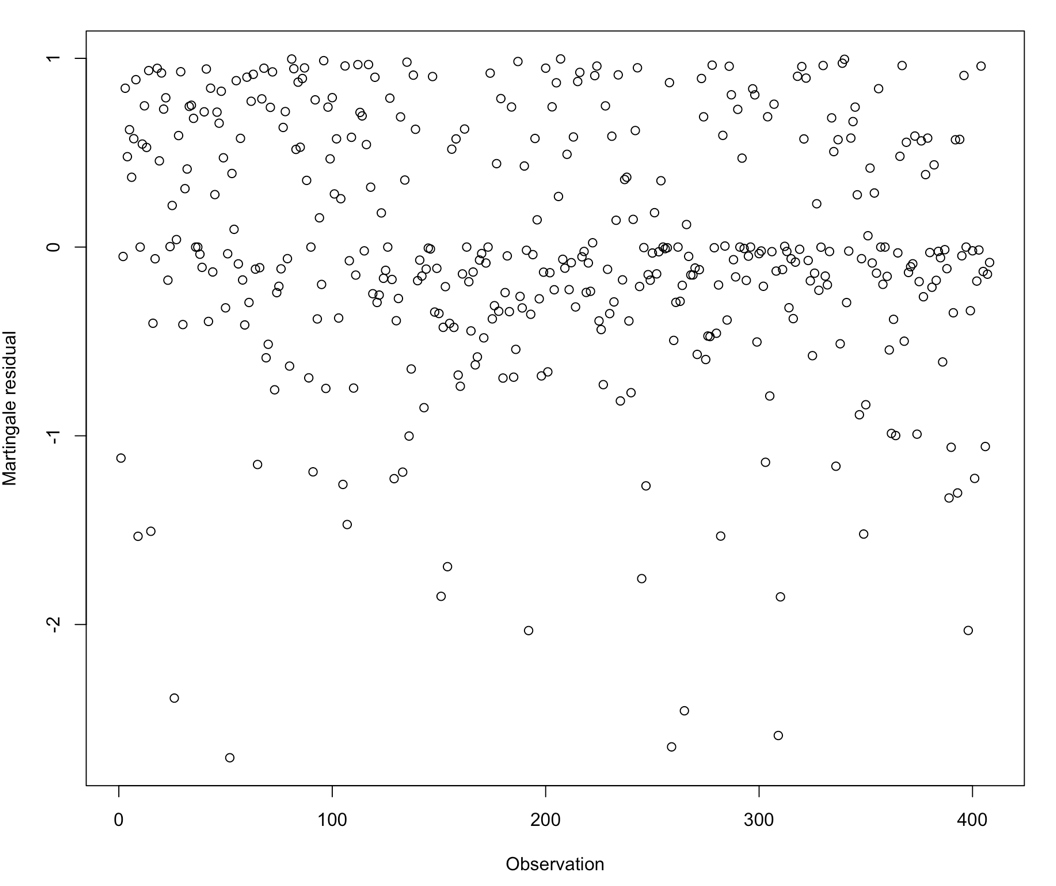

We also evaluated the proportional hazards assumption for each of the covariates in the model that included groups for all four domains at study entry and groups for each of the 6-month changes. The below results to the left are the results reported in the manuscript and to the right are the data for the non-linear term for each covariate. Only one of these p values were <0.05.

|  | **Cox Model** | | | **Proportional Hazards tests** | | |
| --- | --- | --- | --- | --- | --- | --- |
| **Characteristic** | HR | 95% CI | p-value | Chi sq | df | p |
| Baseline memory category 1(ADNI MEM) | 0.22 | 0.11, 0.47 | <0.001 | 3.33 | 1 | 0.068 |
| Baseline memory category 2 (ADNI MEM) | 0.61 | 0.35, 1.07 | 0.085 | 0.35 | 1 | 0.56 |
| Baseline memory category 4 (ADNI MEM) | 1.83 | 1.15, 2.91 | 0.011 | 0.02 | 1 | 0.89 |
| Baseline memory category 5 (ADNI MEM) | 3.12 | 1.94, 5.02 | <0.001 | 2.26 | 1 | 0.13 |
| Baseline executive functioning category 1 (ADNI EFN) | 0.88 | 0.47, 1.64 | 0.69 | 0.00 | 1 | 0.98 |
| Baseline executive functioning category 2 (ADNI EFN) | 1.17 | 0.71, 1.96 | 0.54 | 0.91 | 1 | 0.34 |
| Baseline executive functioning category 4 (ADNI EFN) | 1.16 | 0.72, 1.86 | 0.54 | 0.06 | 1 | 0.80 |
| Baseline executive functioning category 5 (ADNI EFN) | 1.87 | 1.13, 3.07 | 0.014 | 2.60 | 1 | 0.11 |
| Baseline language functioning category 1 (ADNI LAN) | 0.64 | 0.34, 1.21 | 0.17 | 0.78 | 1 | 0.38 |
| Baseline language functioning category 2 (ADNI LAN) | 0.81 | 0.50, 1.32 | 0.40 | 2.97 | 1 | 0.085 |
| Baseline language functioning category 4 (ADNI LAN) | 0.65 | 0.39, 1.07 | 0.093 | 2.29 | 1 | 0.13 |
| Baseline language functioning category 5 (ADNI LAN) | 1.08 | 0.64, 1.83 | 0.76 | 0.43 | 1 | 0.51 |
| Baseline visuospatial functioning category 1 (ADNI VSP) | 0.83 | 0.54, 1.27 | 0.39 | 3.39 | 1 | 0.066 |
| Baseline visuospatial functioning category 4 (ADNI VSP) | 0.63 | 0.37, 1.09 | 0.10 | 0.04 | 1 | 0.85 |
| Baseline visuospatial functioning category 5 (ADNI VSP) | 0.72 | 0.44, 1.17 | 0.18 | 3.72 | 1 | 0.054 |
| Memory score (ADNI MEM) change score category 1 | 0.14 | 0.03, 0.60 | 0.008 | 2.52 | 1 | 0.11 |
| Memory score (ADNI MEM) change score category 2 | 0.47 | 0.29, 0.77 | 0.002 | 0.66 | 1 | 0.42 |
| Memory score (ADNI MEM) change score category 4 | 0.83 | 0.56, 1.24 | 0.37 | 0.66 | 1 | 0.42 |
| Memory score (ADNI MEM) change score category 5 | 1.16 | 0.71, 1.89 | 0.56 | **4.17** | **1** | **0.041** |
| Executive Functioning score (ADNI EFN) change score category 1 | 0.84 | 0.47, 1.50 | 0.56 | 1.52 | 1 | 0.22 |
| Executive Functioning score (ADNI EFN) change score category 2 | 0.93 | 0.60, 1.44 | 0.75 | 0.89 | 1 | 0.35 |
| Executive Functioning score (ADNI EFN) change score category 4 | 1.50 | 0.99, 2.26 | 0.055 | 0.71 | 1 | 0.40 |
| Executive Functioning score (ADNI EFN) change score category 5 | 1.58 | 0.82, 3.07 | 0.17 | 1.92 | 1 | 0.17 |
| Language functioning (ADNI LAN) change score category 1 | 0.72 | 0.35, 1.48 | 0.37 | 1.34 | 1 | 0.25 |
| Language functioning (ADNI LAN) change score category 2 | 0.56 | 0.36, 0.89 | 0.013 | 0.04 | 1 | 0.85 |
| Language functioning (ADNI LAN) change score category 4 | 0.88 | 0.60, 1.29 | 0.51 | 0.44 | 1 | 0.51 |
| Language functioning (ADNI LAN) change score category 5 | 0.88 | 0.49, 1.56 | 0.65 | 0.11 | 1 | 0.75 |
| Visuospatial functioning (ADNI VSP) change score category 1 | 1.80 | 0.81, 3.99 | 0.15 | 0.02 | 1 | 0.90 |
| Visuospatial functioning (ADNI VSP) change score category 2 | 0.96 | 0.60, 1.52 | 0.85 | 0.17 | 1 | 0.68 |
| Visuospatial functioning (ADNI VSP) change score category 4 | 0.89 | 0.59, 1.34 | 0.57 | 0.23 | 1 | 0.63 |
| Visuospatial functioning (ADNI VSP) change score category 5 | 1.34 | 0.73, 2.46 | 0.34 | 0.00 | 1 | 0.98 |
| Age | 1.01 | 0.99, 1.04 | 0.35 | 2.44 | 1 | 0.12 |
| Gender | 1.78 | 1.29, 2.46 | <0.001 | 3.12 | 1 | 0.077 |
| Educational attainment | 1.04 | 0.98, 1.10 | 0.22 | 0.51 | 1 | 0.48 |
| Global test, all covariates |  |  |  | 41.80 | 34 | 0.17 |

Note: Abbreviations: CI = Confidence Interval, HR = Hazard Ratio

### Supplemental Results 2. Model checking for the entire cohort.

We performed additional analyses to check the proportional hazards assumption. Each variable is added to the model with a non-linear term. The p value column in this table shows results of tests of whether that non-linear term is statistically different from the null. The findings for categories defined by memory levels at enrollment had a p value <0.05, as did the categories defined by visuospatial at enrollment. The nonlinear terms for categories defined by changes in domain scores for all of the domains (including memory and visuospatial) all had p values >0.05.

| **Covariate** | **Chi-square** | **df** | **p** |
| --- | --- | --- | --- |
| Baseline memory category (ADNI MEM) | 11.82 | 4 | 0.02 |
| Baseline executive functioning category (ADNI EFN) | 3.23 | 4 | 0.52 |
| Baseline language functioning category (ADNI LAN) | 4.90 | 4 | 0.30 |
| Baseline visuospatial functioning category (ADNI VSP) | 8.76 | 3 | 0.03 |
| Memory score (ADNI MEM) change score categories | 8.39 | 4 | 0.08 |
| Executive Functioning score (ADNI EFN) change score categories | 8.01 | 4 | 0.09 |
| Language functioning (ADNI LAN) change score categories | 3.32 | 4 | 0.51 |
| Visuospatial functioning (ADNI VSP) change score categories | 1.46 | 4 | 0.83 |
| Age | 3.78 | 1 | 0.052 |
| Gender | 0.20 | 1 | 0.65 |
| Educational attainment | 2.90 | 1 | 0.09 |
| Global test, all covariates | 52.34 | 34 | 0.02 |

We performed additional analyses of the memory scores at enrollment. There were few people in the highest category who subsequently converted from MCI at the month 6 visit to AD dementia. Those who did had a lot of follow-up time. There were some people who did not convert who only had shorter follow-up time, but there were any who did not convert who had follow-up time that was similar to those who did convert, as seen in the following figure:

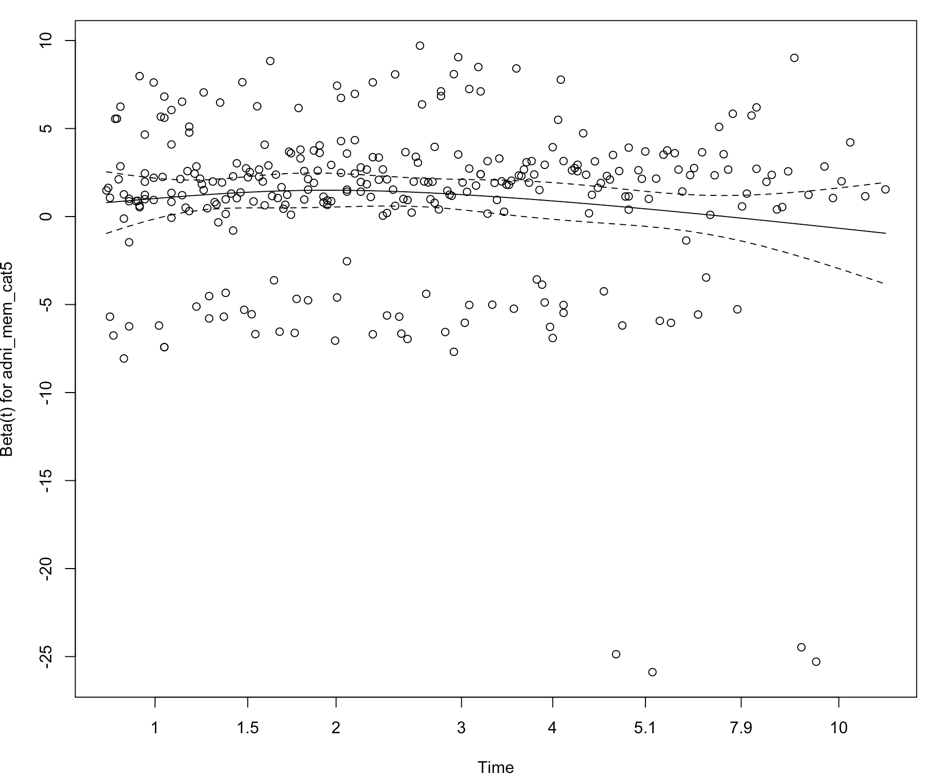

We also evaluated Martingale residuals and found no particular outliers:

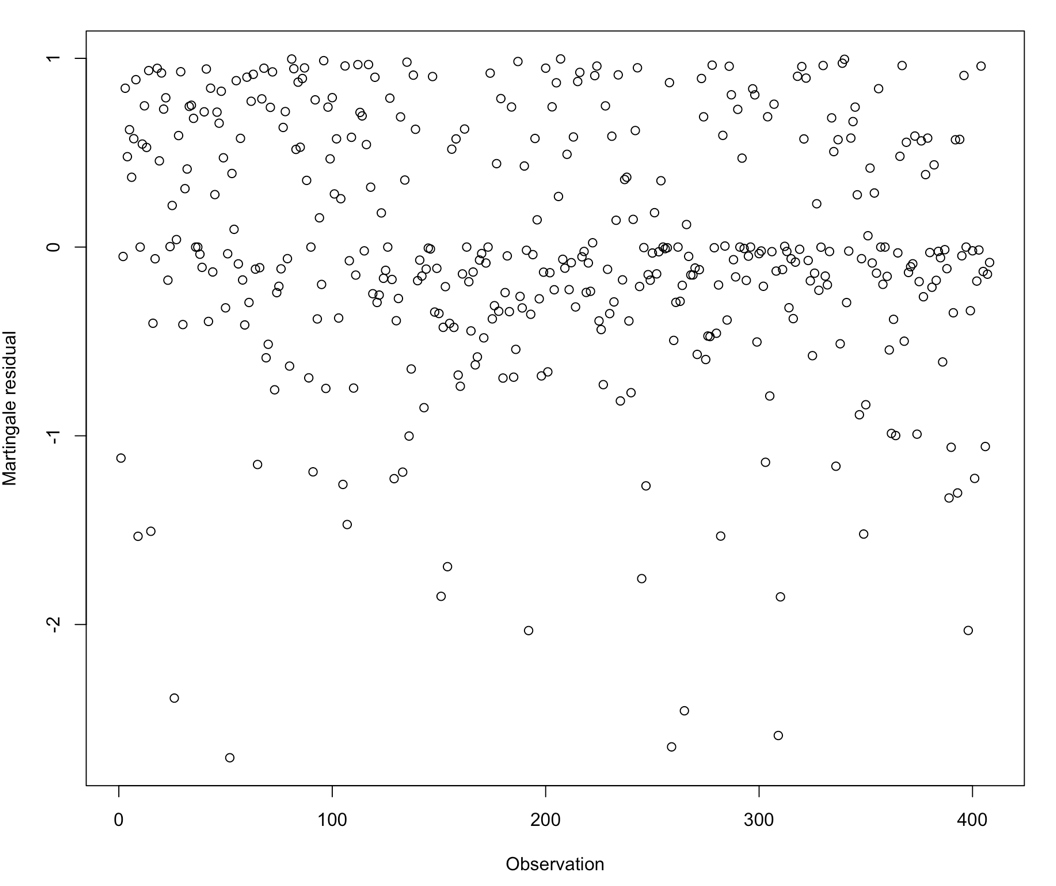

We also evaluated the proportional hazards assumption for each of the covariates in the model that included groups for all four domains at study entry and groups for each of the 6-month changes. The below results to the left are the results reported in the manuscript and to the right are the data for the non-linear term for each covariate. None of these p values were <0.05.

|  | **Cox Model** | | | **Proportional Hazards tests** | | |
| --- | --- | --- | --- | --- | --- | --- |
| **Characteristic** | HR | 95% CI | p-value | Chi sq | df | p |
| Baseline memory category 1(ADNI MEM) | 0.20 | 0.11, 0.36 | <0.001 | **10.22** | **1** | **0.001** |
| Baseline memory category 2 (ADNI MEM) | 0.55 | 0.36, 0.84 | 0.005 | 0.08 | 1 | 0.78 |
| Baseline memory category 4 (ADNI MEM) | 1.65 | 1.17, 2.32 | 0.004 | 1.31 | 1 | 0.25 |
| Baseline memory category 5 (ADNI MEM) | 2.80 | 1.95, 4.02 | <0.001 | 1.35 | 1 | 0.25 |
| Baseline executive functioning category 1 (ADNI EFN) | 0.67 | 0.42, 1.06 | 0.090 | 0.03 | 1 | 0.86 |
| Baseline executive functioning category 2 (ADNI EFN) | 0.93 | 0.63, 1.39 | 0.73 | 0.55 | 1 | 0.46 |
| Baseline executive functioning category 4 (ADNI EFN) | 1.15 | 0.80, 1.64 | 0.46 | 0.63 | 1 | 0.43 |
| Baseline executive functioning category 5 (ADNI EFN) | 1.83 | 1.25, 2.68 | 0.002 | 2.95 | 1 | 0.086 |
| Baseline language functioning category 1 (ADNI LAN) | 0.63 | 0.39, 1.03 | 0.065 | 2.35 | 1 | 0.12 |
| Baseline language functioning category 2 (ADNI LAN) | 0.90 | 0.61, 1.33 | 0.60 | 1.21 | 1 | 0.27 |
| Baseline language functioning category 4 (ADNI LAN) | 0.76 | 0.52, 1.11 | 0.16 | 0.24 | 1 | 0.63 |
| Baseline language functioning category 5 (ADNI LAN) | 1.00 | 0.67, 1.48 | 0.98 | 2.11 | 1 | 0.15 |
| Baseline visuospatial functioning category 1 (ADNI VSP) | 0.88 | 0.63, 1.22 | 0.45 | 1.91 | 1 | 0.17 |
| Baseline visuospatial functioning category 4 (ADNI VSP) | 0.99 | 0.67, 1.47 | 0.98 | 2.22 | 1 | 0.14 |
| Baseline visuospatial functioning category 5 (ADNI VSP) | 0.83 | 0.57, 1.21 | 0.33 | **6.88** | **1** | **0.009** |
| Memory score (ADNI MEM) change score category 1 | 0.29 | 0.11, 0.82 | 0.019 | **6.64** | **1** | **0.010** |
| Memory score (ADNI MEM) change score category 2 | 0.72 | 0.50, 1.04 | 0.076 | 0.17 | 1 | 0.68 |
| Memory score (ADNI MEM) change score category 4 | 1.03 | 0.77, 1.37 | 0.83 | 0.04 | 1 | 0.84 |
| Memory score (ADNI MEM) change score category 5 | 1.29 | 0.89, 1.88 | 0.18 | 1.76 | 1 | 0.18 |
| Executive Functioning score (ADNI EFN) change score category 1 | 0.64 | 0.40, 1.04 | 0.070 | **4.95** | **1** | **0.026** |
| Executive Functioning score (ADNI EFN) change score category 2 | 0.87 | 0.64, 1.19 | 0.39 | 0.12 | 1 | 0.73 |
| Executive Functioning score (ADNI EFN) change score category 4 | 1.05 | 0.76, 1.44 | 0.76 | 0.78 | 1 | 0.38 |
| Executive Functioning score (ADNI EFN) change score category 5 | 1.32 | 0.79, 2.19 | 0.29 | 0.32 | 1 | 0.57 |
| Language functioning (ADNI LAN) change score category 1 | 0.74 | 0.42, 1.30 | 0.29 | 2.39 | 1 | 0.12 |
| Language functioning (ADNI LAN) change score category 2 | 0.63 | 0.45, 0.88 | 0.007 | 0.00 | 1 | 0.98 |
| Language functioning (ADNI LAN) change score category 4 | 1.02 | 0.75, 1.38 | 0.90 | 1.26 | 1 | 0.26 |
| Language functioning (ADNI LAN) change score category 5 | 1.23 | 0.79, 1.91 | 0.37 | 0.01 | 1 | 0.91 |
| Visuospatial functioning (ADNI VSP) change score category 1 | 1.37 | 0.71, 2.64 | 0.35 | 0.07 | 1 | 0.79 |
| Visuospatial functioning (ADNI VSP) change score category 2 | 0.76 | 0.54, 1.06 | 0.11 | 0.00 | 1 | 0.97 |
| Visuospatial functioning (ADNI VSP) change score category 4 | 0.90 | 0.65, 1.24 | 0.51 | 0.00 | 1 | 0.96 |
| Visuospatial functioning (ADNI VSP) change score category 5 | 0.85 | 0.51, 1.42 | 0.54 | 1.21 | 1 | 0.27 |
| Age | 1.01 | 0.99, 1.02 | 0.50 | 3.78 | 1 | 0.052 |
| Gender | 1.51 | 1.18, 1.94 | 0.001 | 0.20 | 1 | 0.65 |
| Educational attainment | 1.06 | 1.02, 1.11 | 0.005 | 2.90 | 1 | 0.089 |
| Global test, all covariates |  |  |  | **52.34** | **34** | **0.023** |

Note: Abbreviations: CI = Confidence Interval, HR = Hazard Ratio

### Supplemental Table S6. Cox model results for memory at baseline and 6-month change (as in Table 2, left columns) plus each of *APOE* genotype, Cortical Gray Matter Volume, and Hippocampal Volume

|  | **Memory alone** | | ***APOE* genotype** | | **Cortical gray matter** | | **Hippocampal volume** | |
| --- | --- | --- | --- | --- | --- | --- | --- | --- |
|  | **All**  **(n = 764)** | **Amyloid + (n=408)** | **All**  **(n = 764)** | **Amyloid + (n=408)** | **All**  **(n = 764)** | **Amyloid + (n=408)** | **All**  **(n = 764)** | **Amyloid + (n=408)** |
| **6-month change categories** |  |  |  |  |  |  |  |  |
| Improved a lot HR, 95% CI | **0.24 (0.09, 0.66)** | **0.16 (0.04, 0.65)** | **0.26 (0.09, 0.71)** | **0.16 (0.04, 0.65)** | **0.24 (0.08, 0.76)** | **0.18 (0.04, 0.73)** | **0.27 (0.08, 0.86)** | **0.20 (0.05, 0.82)** |
| Improved a little HR, 95% CI | 0.74 (0.53, 1.03) | **0.56 (0.37, 0.86)** | 0.72 (0.51, 1.00) | **0.56 (0.36, 0.86)** | 0.77 (0.55, 1.09) | **0.63 (0.41, 0.97)** | 0.80 (0.57, 1.13) | **0.63 (0.41, 0.98)** |
| Minimal change (reference) | 1 (reference) | 1 (reference) | 1 (reference) | 1 (reference) | 1 (reference) | 1 (reference) | 1 (reference) | 1 (reference) |
| A little worse HR, 95% CI | 1.10 (0.83, 1.45) | 0.97 (0.68, 1.39) | 1.07 (0.81, 1.42) | 0.97 (0.68, 1.39) | 1.03 (0.78, 1.37) | 0.93 (0.64, 1.35) | 1.04 (0.78, 1.39) | 0.91 (0.63, 1.32) |
| A lot worse HR, 95% CI | 1.39 (0.96, 2.00) | 1.24 (0.77, 2.00) | **1.46 (1.01, 2.10)** | 1.26 (0.79, 2.03) | 1.38 (0.94, 2.02) | 1.27 (0.78, 2.08) | 1.43 (0.98, 2.10) | 1.20 (0.73, 1.98) |
| **Baseline value categories** |  |  |  |  |  |  |  |  |
| High HR, 95% CI | **0.17 (0.10, 0.29)** | **0.20 (0.10, 0.39)** | **0.18 (0.10, 0.31)** | **0.20 (0.10, 0.40)** | **0.16 (0.09, 0.29)** | **0.20 (0.09, 0.44)** | **0.17 (0.09, 0.32)** | **0.21 (0.10, 0.45)** |
| Medium high HR, 95% CI | **0.49 (0.32, 0.73)** | **0.49 (0.29, 0.82)** | **0.47 (0.31, 0.71)** | **0.53 (0.32, 0.89)** | **0.50 (0.33, 0.76)** | **0.56 (0.33, 0.96)** | **0.51 (0.34, 0.78)** | **0.54 (0.32, 0.92)** |
| Medium (reference) | 1 (reference) | 1 (reference) | 1 (reference) | 1 (reference) | 1 (reference) | 1 (reference) | 1 (reference) | 1 (reference) |
| Medium low HR, 95% CI | **1.82 (1.30, 2.54)** | **1.66 (1.10, 2.52)** | **1.71 (1.22, 2.39)** | **1.64 (1.07, 2.51)** | **1.79 (1.27, 2.51)** | **1.70 (1.10, 2.63)** | **1.70 (1.21, 2.38)** | **1.61 (1.04, 2.50)** |
| Low HR, 95% CI | **3.60 (2.61, 4.97)** | **2.85 (2.61, 4.97)** | **3.34 (2.41, 4.61)** | **3.20 (2.14, 4.78)** | **3.43 (2.47, 4.76)** | **3.39 (2.24, 5.12)** | **3.13 (2.25, 4.36)** | **3.08 (2.04, 4.67)** |
| **Demographic characteristics** |  |  |  |  |  |  |  |  |
| Age HR, 95% CI | 1.02 (1.00, 1.03) | 1.01 (0.99, 1.03) | **1.02 (1.00, 1.04)** | 1.02 (1.00, 1.04) | 1.00 (0.98, 1.02) | 1.00 (0.98, 1.03) | 1.00 (0.98, 1.02) | 1.00 (0.98, 1.03) |
| Female sex HR, 95% CI | **1.44 (1.13, 1.82)** | **1.78 (1.30, 2.44)** | **1.49 (1.16, 1.90)** | **1.72 (1.25, 2.37)** | **1.35 (1.05, 1.74)** | **1.55 (1.10, 2.18)** | **1.32 (1.02, 1.71)** | **1.62 (1.16, 2.25)** |
| Education HR, 95% CI | 1.03 (0.99, 1.07) | 1.03 (0.97, 1.08) | 1.04 (1.00, 1.09) | 1.03 (0.97, 1.08) | 1.03 (0.99, 1.08) | 1.02 (0.96, 1.08) | 1.03 (0.99, 1.08) | 1.02 (0.97, 1.08) |
| **Additional covariates** |  |  |  |  |  |  |  |  |
| ≥1 *APOE* ε4 allele HR, 95% CI |  |  | **2.03 (1.59, 2.60)** | **1.48 (1.08, 2.03)** |  |  |  |  |
| Cortical gray matter |  |  |  |  | **0.60 (0.47, 0.76)** | **0.67 (0.49, 0.91)** |  |  |
| Hippocampal vol. HR, 95% CI |  |  |  |  |  |  | **0.54 (0.41, 0.71)** | **0.57 (0.39, 0.84)** |

Note: Abbreviations: CI = Confidence Interval, HR = Hazard Ratio

### Supplemental Table S7. Cox model results for memory at baseline and 6-month change (as in Table 2, left columns) plus each of Entorhinal thickness, Temporal lobe thickness, and Medial temporal lobe thickness

|  | **Memory alone** | | **Entorhinal thickness** | | **Temporal thickness** | | **Medial temporal lobe thickness** | |
| --- | --- | --- | --- | --- | --- | --- | --- | --- |
|  | **All**  **(n = 764)** | **Amyloid + (n=408)** | **All**  **(n = 764)** | **Amyloid + (n=408)** | **All**  **(n = 764)** | **Amyloid + (n=408)** | **All**  **(n = 764)** | **Amyloid + (n=408)** |
| **6-month change categories** |  |  |  |  |  |  |  |  |
| Improved a lot HR, 95% CI | **0.24 (0.09, 0.66)** | **0.16 (0.04, 0.65)** | **0.25 (0.08, 0.80)** | **0.19 (0.05, 0.80)** | **0.25 (0.08, 0.80)** | **0.19 (0.05, 0.81)** | **0.25 (0.08, 0.80)** | **0.19 (0.05, 0.80)** |
| Improved a little HR, 95% CI | 0.74 (0.53, 1.03) | **0.56 (0.37, 0.86)** | 0.72 (0.51, 1.01) | **0.61 (0.40, 0.95)** | 0.72 (0.51, 1.02) | **0.63 (0.41, 0.98)** | 0.72 (0.51, 1.02) | **0.62 (0.40, 0.96)** |
| Minimal change (reference) | 1 (reference) | 1 (reference) | 1 (reference) | 1 (reference) | 1 (reference) | 1 (reference) | 1 (reference) | 1 (reference) |
| A little worse HR, 95% CI | 1.10 (0.83, 1.45) | 0.97 (0.68, 1.39) | 1.01 (0.76, 1.34) | 0.90 (0.62, 1.31) | 0.97 (0.73, 1.29) | 0.82 (0.56, 1.20) | 0.98 (0.73, 1.30) | 0.84 (0.58, 1.23) |
| A lot worse HR, 95% CI | 1.39 (0.96, 2.00) | 1.24 (0.77, 2.00) | 1.31 (0.89, 1.92) | 1.24 (0.75, 2.04) | 1.23 (0.84, 1.81) | 1.06 (0.64, 1.75) | 1.26 (0.85, 1.85) | 1.13 (0.68, 1.87) |
| **Baseline value categories** |  |  |  |  |  |  |  |  |
| High HR, 95% CI | **0.17 (0.10, 0.29)** | **0.20 (0.10, 0.39)** | **0.16 (0.09, 0.30)** | **0.21 (0.10, 0.47)** | **0.16 (0.09, 0.30** | **0.23 (0.11, 0.50)** | **0.16 (0.09. 0.30)** | **0.22 (0.10, 0.49)** |
| Medium high HR, 95% CI | **0.49 (0.32, 0.73)** | **0.49 (0.29, 0.82)** | **0.51 (0.33, 0.77)** | **0.56 (0.33, 0.96)** | **0.52 (0.34, 0.79)** | 0.61 (0.36, 1.05) | **0.51 (0.34, 0.78)** | 0.59 (0.34, 1.00) |
| Medium (reference) | 1 (reference) | 1 (reference) | 1 (reference) | 1 (reference) | 1 (reference) | 1 (reference) | 1 (reference) | 1 (reference) |
| Medium low HR, 95% CI | **1.82 (1.30, 2.54)** | **1.66 (1.10, 2.52)** | **1.73 (1.23, 2.42)** | **1.71 (1.11, 2.64)** | **1.80 (1.28, 2.53)** | **1.80 (1.16, 2.79)** | **1.81 (1.29, 2.54)** | **1.80 (1.16, 2.79)** |
| Low HR, 95% CI | **3.60 (2.61, 4.97)** | **2.85 (2.61, 4.97)** | **3.17 (2.28, 4.42)** | **3.14 (2.08, 4.76)** | **3.13 (2.25, 4.36)** | **3.22 (2.13, 4.87)** | **3.21 (2.31, 4.47)** | **3.22 (2.13, 4.86)** |
| **Demographic characteristics** |  |  |  |  |  |  |  |  |
| Age HR, 95% CI | 1.02 (1.00, 1.03) | 1.01 (0.99, 1.03) | 1.01 (0.99, 1.02) | 1.01 (0.99, 1.03) | 1.00 (0.98, 1.01) | 1.00 (0.98, 1.02) | 1.00 (0.99, 1.02) | 1.01 (0.98, 1.03) |
| Female sex HR, 95% CI | **1.44 (1.13, 1.82)** | **1.78 (1.30, 2.44)** | **1.43 (1.11, 1.84)** | **1.74 (1.26, 2.41)** | **1.49 (1.16, 1.91)** | **1.71 (1.24, 2.36)** | **1.47 (1.14, 1.89)** | **1.77 (1.28, 2.44)** |
| Education HR, 95% CI | 1.03 (0.99, 1.07) | 1.03 (0.97, 1.08) | 1,03 (0.99, 1.08) | 1.02 (0.97, 1.08) | 1.02 (0.98, 1.07) | 1.01 (0.95, 1.07) | 1.03 (0.99, 1.07) | 1.02 (0.97, 1.08) |
| **Additional covariates** |  |  |  |  |  |  |  |  |
| ER thickness HR, 95% CI |  |  | **0.69 (0.54, 0.87)** | 0.73 (0.52, 1.03) |  |  |  |  |
| Temp thickness HR, 95% CI |  |  |  |  | **0.47 (0.36, 0.61)** | **0.47 (0.33, 0.67)** |  |  |
| M temp thickness HR, 95% CI |  |  |  |  |  |  | **0.59 (0.46, 0.76)** | **0.57 (0.40, 0.80)** |

Note: Abbreviations: CI = Confidence Interval, HR = Hazard Ratio

### Supplemental Table S8. Cox model results for memory at baseline and 6-month change (as in Table 2, left columns) plus each of Lateral Temporal Lobe Thickness, Frontal Lobe Thickness, and Cingulate Thickness

|  | **Memory alone** | | **Lateral Temporal Lobe Thickness** | | **Frontal lobe thickness** | | **Cingulate Thickness** | |
| --- | --- | --- | --- | --- | --- | --- | --- | --- |
|  | **All**  **(n = 764)** | **Amyloid + (n=408)** | **All**  **(n = 764)** | **Amyloid + (n=408)** | **All**  **(n = 764)** | **Amyloid + (n=408)** | **All**  **(n = 764)** | **Amyloid + (n=408)** |
| **6-month change categories** |  |  |  |  |  |  |  |  |
| Improved a lot HR, 95% CI | **0.24 (0.09, 0.66)** | **0.16 (0.04, 0.65)** | **0.25 (0.08, 0.80)** | **0.20 (0.05, 0.82)** | **0.25 (0.08, 0.81)** | **0.20 (0.05, 0.82)** | **0.25 (0.08, 0.81)** | **0.19 (0.05, 0.77)** |
| Improved a little HR, 95% CI | 0.74 (0.53, 1.03) | **0.56 (0.37, 0.86)** | 0.74 (0.52, 1.04) | **0.64 (0.42, 0.99)** | 0.75 (0.53, 1.05) | **0.61 (0.39, 0.93)** | 0.75 (0.54, 1.06) | **0.59 (0.38, 0.91)** |
| Minimal change (reference) | 1 (reference) | 1 (reference) | 1 (reference) | 1 (reference) | 1 (reference) | 1 (reference) | 1 (reference) | 1 (reference) |
| A little worse HR, 95% CI | 1.10 (0.83, 1.45) | 0.97 (0.68, 1.39) | 0.99 (0.74, 1.31) | 0.85 (0.58, 1.23) | 1.05 (0.79, 1.40) | 0.91 (0.63, 1.32) | 1.02 (0.76, 1.36) | 0.92 (0.63, 1.33) |
| A lot worse HR, 95% CI | 1.39 (0.96, 2.00) | 1.24 (0.77, 2.00) | 1.26 (0.86, 1.85) | 1.06 (0.64, 1.76) | 1.38 (0.94, 2.03) | 1.27 (0.78, 2.08) | 1.39 (0.94, 2.04) | 1.26 (0.76, 2.07) |
| **Baseline value categories** |  |  |  |  |  |  |  |  |
| High HR, 95% CI | **0.17 (0.10, 0.29)** | **0.20 (0.10, 0.39)** | **0.16 (0.08, 0.29)** | **0.22 (0.10, 0.48)** | **0.15 (0.08, 0.28)** | **0.19 (0.09. 0.42)** | **0.14 (0.08, 0.26)** | **0.18 (0.08, 0.40)** |
| Medium high HR, 95% CI | **0.49 (0.32, 0.73)** | **0.49 (0.29, 0.82)** | **0.50 (0.33, 0.76)** | **0.61 (0.36, 1.05)** | **0.47 (0.31, 0.71)** | **0.54 (0.32, 0.92)** | **0.45 (0.30, 0.69)** | **0.51 (0.30, 0.88)** |
| Medium (reference) | 1 (reference) | 1 (reference) | 1 (reference) | 1 (reference) | 1 (reference) | 1 (reference) | 1 (reference) | 1 (reference) |
| Medium low HR, 95% CI | **1.82 (1.30, 2.54)** | **1.66 (1.10, 2.52)** | **1.77 (1.26, 2.48)** | **1.75 (1.13, 2.72)** | **1.79 (1.28, 2.52)** | **1.72 (1.11, 2.66)** | **1.75 (1.24, 2.45)** | **1.71 (1.11, 2.64)** |
| Low HR, 95% CI | **3.60 (2.61, 4.97)** | **2.85 (2.61, 4.97)** | **3.13 (2.25, 4.36)** | **3.24 (2.14, 4.90)** | **3.39 (2.44, 4.71)** | **3.27 (2.17, 4.94)** | **3.23 (2.32, 4.50)** | **3.12 (2.05, 4.74)** |
| **Demographic characteristics** |  |  |  |  |  |  |  |  |
| Age HR, 95% CI | 1.02 (1.00, 1.03) | 1.01 (0.99, 1.03) | 1.00 (0.98, 1.01) | 1.00 (0.98, 1.02) | 1.00 (0.99, 1.02) | 1.01 (0.99, 1.03) | 1.01 (1.00, 1.03) | 1.02 (0.99, 1.04) |
| Female sex HR, 95% CI | **1.44 (1.13, 1.82)** | **1.78 (1.30, 2.44)** | **1.50 (1.17, 1.93)** | **1.67 (1.21, 2.31)** | **1.56 (1.21, 2.00)** | **1.79 (1.30, 2.48)** | **1.55 (1.21, 2.00)** | **1.83 (1.32, 2.54)** |
| Education HR, 95% CI | 1.03 (0.99, 1.07) | 1.03 (0.97, 1.08) | 1.02 (0.98, 1.07) | 1.00 (0.95, 1.06) | 1.04 (1.00 1.08) | 1.02 (0.96, 1.08) | 1.04 (1.00, 1.08) | 1.03 (0.97, 1.09) |
| **Additional covariates** |  |  |  |  |  |  |  |  |
| L Temp Thickness HR, 95% CI |  |  | **0.44 (0.34, 0.58)** | **0.46 (0.33, 0.65)** |  |  |  |  |
| Frontal Thickness HR, 95% CI |  |  |  |  | **0.66 (0.51, 0.85)** | **0.68 (0.49, 0.94)** |  |  |
| Cing. Thickness HR, 95% CI |  |  |  |  |  |  | **0.76 (0.59, 0.96)** | 0.81 (0.59, 1.11) |

Note: Abbreviations: CI = Confidence Interval, HR = Hazard Ratio

### Supplemental Table S9. Cox model results for memory at baseline and 6-month change (as in Table 2, left columns) plus each of Parietal Lobe Thickness, Occipital Lobe Thickness, and Sensory / Motor Cortical Thickness

|  | **Memory alone** | | **Parietal lobe thickness** | | **Occipital lobe thickness** | | **Sensory / Motor Cortical Thickness** | |
| --- | --- | --- | --- | --- | --- | --- | --- | --- |
|  | **All**  **(n = 764)** | **Amyloid + (n=408)** | **All**  **(n = 764)** | **Amyloid + (n=408)** | **All**  **(n = 764)** | **Amyloid + (n=408)** | **All**  **(n = 764)** | **Amyloid + (n=408)** |
| **6-month change categories** |  |  |  |  |  |  |  |  |
| Improved a lot HR, 95% CI | **0.24 (0.09, 0.66)** | **0.16 (0.04, 0.65)** | **0.25 (0.08, 0.79)** | **0.19 (0.05, 0.80)** | **0.24 (0.07, 0.75)** | **0.18 (0.04, 0.73)** | **0.24 (0.08, 0.76)** | **0.18 (0.04, 0.75)** |
| Improved a little HR, 95% CI | 0.74 (0.53, 1.03) | **0.56 (0.37, 0.86)** | 0.75 (0.54, 1.06) | **0.61 (0.40, 0.95)** | 0.74 (0.52, 1.04) | **0.60 (0.39, 0.92)** | 0.74 (0.53, 1.04) | **0.59 (0.38, 0.91)** |
| Minimal change (reference) | 1 (reference) | 1 (reference) | 1 (reference) | 1 (reference) | 1 (reference) | 1 (reference) | 1 (reference) | 1 (reference) |
| A little worse HR, 95% CI | 1.10 (0.83, 1.45) | 0.97 (0.68, 1.39) | 1.05 (0.79, 1.39) | 0.91 (0.63, 1.32) | 1.04 (0.78, 1.38) | 0.92 (0.63, 1.34) | 1.05 (0.79, 1.40) | 0.93 (0.64, 1.35) |
| A lot worse HR, 95% CI | 1.39 (0.96, 2.00) | 1.24 (0.77, 2.00) | **1.36 (0.93, 1.99)** | 1.21 (0.73, 1.99) | 1.37 (0.93, 2.01) | 1.25 (0.76, 2.06) | 1.36 (0.93, 2.01) | 1.28 (0.77, 2.10) |
| **Baseline value categories** |  |  |  |  |  |  |  |  |
| High HR, 95% CI | **0.17 (0.10, 0.29)** | **0.20 (0.10, 0.39)** | **0.16 (0.09, 0.29)** | **0.21 (0.09, 0.45)** | **0.15 (0.08, 0.28)** | **0.20 (0.09, 0.43)** | **0.15 (0.08, 0.27)** | **0.19 (0.09, 0.41)** |
| Medium high HR, 95% CI | **0.49 (0.32, 0.73)** | **0.49 (0.29, 0.82)** | **0.47 (0.31, 0.71)** | **0.54 (0.32, 0.93)** | **0.48 (0.31, 0.73)** | **0.55 (0.32, 0.93)** | **0.47 (0.31, 0.71)** | **0.54 (0.31, 0.91)** |
| Medium (reference) | 1 (reference) | 1 (reference) | 1 (reference) | 1 (reference) | 1 (reference) | 1 (reference) | 1 (reference) | 1 (reference) |
| Medium low HR, 95% CI | **1.82 (1.30, 2.54)** | **1.66 (1.10, 2.52)** | **1.70 (1.21, 2.38)** | **1.59 (1.02, 2.47)** | **1.78 (1.27, 2.50)** | **1.70 (1.10, 2.63)** | **1.77 (1.26, 2.48)** | **1.71 (1.11, 2.64)** |
| Low HR, 95% CI | **3.60 (2.61, 4.97)** | **2.85 (2.61, 4.97)** | **3.09 (2.22, 4.31)** | **3.01 (1.99, 4.57)** | **3.40 (2.45, 4.73)** | **3.29 (2.18, 4.97)** | **3.42 (2.46, 4.76)** | **3.32 (2.20, 5.01)** |
| **Demographic characteristics** |  |  |  |  |  |  |  |  |
| Age HR, 95% CI | 1.02 (1.00, 1.03) | 1.01 (0.99, 1.03) | 1.00 (0.99, 1.02) | 1.01 (0.99, 1.03) | 1.01 (0.99, 1.02) | 1.01 (0.99, 1.03) | 1.01 (0.99, 1.02) | 1.01 (0.99, 1.03) |
| Female sex HR, 95% CI | **1.44 (1.13, 1.82)** | **1.78 (1.30, 2.44)** | **1.51 (1.18, 1.94)** | **1.67 (1.20, 2.31)** | **1.51 (1.17, 1.94)** | **1.75 (1.27, 2.43)** | **1.54 (1.20, 1.98)** | **1.78 (1.29, 2.47)** |
| Education HR, 95% CI | 1.03 (0.99, 1.07) | 1.03 (0.97, 1.08) | 1.03 (0.99, 1.08) | 1.01 (0.96, 1.07) | 1.04 (1.00, 1.08) | 1.02 (0.97, 1.08) | 1.04 (1.00, 1.08) | 1.02 (0.97, 1.08) |
| **Additional covariates** |  |  |  |  |  |  |  |  |
| Parietal thickness HR, 95% CI |  |  | **0.58 (0.45, 0.76)** | **0.57 (0.47, 0.79)** |  |  |  |  |
| Occip thickness HR, 95% CI |  |  |  |  | 0.81 (0.64, 1.04) | 0.76 (0.56, 1.04) |  |  |
| Sen/Mot thickness HR, 95% CI |  |  |  |  |  |  | 0.84 (0.65, 1.09) | 0.87 (0.64, 1.19) |

Note: Abbreviations: CI = Confidence Interval, HR = Hazard Ratio

### Supplemental Table S10. Cox model results for executive functioning at baseline and 6-month change (as in Table 2, left columns) plus each of *APOE* genotype, Cortical Gray Matter Volume, and Hippocampal Volume

|  | **Executive Functioning** | | ***APOE* genotype** | | **Cortical gray matter** | | **Hippocampal volume** | |
| --- | --- | --- | --- | --- | --- | --- | --- | --- |
|  | **All**  **(n = 764)** | **Amyloid + (n=408)** | **All**  **(n = 764)** | **Amyloid + (n=408)** | **All**  **(n = 764)** | **Amyloid + (n=408)** | **All**  **(n = 764)** | **Amyloid + (n=408)** |
| **6 month change categories** |  |  |  |  |  |  |  |  |
| Improved a lot HR, 95% CI | **0.49 (0.31, 0.79)** | **0.57 (0.33, 0.98)** | **0.50 (0.32, 0.80)** | 0.58 (0.33, 1.00) | **0.49 (0.30, 0.79)** | 0.61 (0.35, 1.08) | **0.49 (0.31, 0.80)** | 0.63 (0.36, 1.12) |
| Improved a little HR, 95% CI | **0.74 (0.55, 1.00)** | 0.80 (0.54, 1.19) | **0.70 (0.52, 0.95)** | 0.77 (0.52, 1.15) | **0.71 (0.53, 0.97)** | **0.80 (0.53, 1.20)** | 0.75 (0.56, 1.02) | 0.81 (0.54, 1.22) |
| Minimal change (reference) | 1 (reference) | 1 (reference) | 1 (reference) | 1 (reference) | 1 (reference) | 1 (reference) | 1 (reference) | 1 (reference) |
| A little worse HR, 95% CI | 1.17 (0.87, 1.58) | **1.56 (1.07, 2.29)** | 1.15 (0.85, 1.55) | **1.56 (1.07, 2.28)** | 1.04 (0.76, 1.42) | 1.43 (0.96, 2.14) | 1.10 (0.81, 1.50) | 1.45 (0.98, 2.16) |
| A lot worse HR, 95% CI | 1.39 (0.86, 2.26) | 1.76 (0.94, 3.29) | 1.39 (0.85, 2.25) | 1.76 (0.95, 3.29) | 1.35 (0.83, 2.20) | 1.81 (0.97, 3.39) | 1.48 (0.91. 2.40) | **2.09 (1.11, 3.92)** |
| **Baseline values** |  |  |  |  |  |  |  |  |
| High HR, 95% CI | **0.37 (0.24, 0.57)** | **0.37 (0.21, 0.66)** | **0.40 (0.26, 0.62)** | **0.37 (0.20, 0.66)** | **0.37 (0.24, 0.59)** | **0.37 (0.20, 0.68)** | **0.39 (0.25, 0.62)** | **0.40 (0.21, 0.74)** |
| Medium high HR, 95% CI | 0.69 (0.47, 1.01) | 0.71 (0.44, 1.15) | **0.65 (0.45, 0.96)** | 0.65 (0.40, 1.05) | 0.70 (0.47, 1.04) | 0.77 (0.47, 1.27) | 0.76 (0.51, 1.12) | 0.94 (0.57, 1.56) |
| Medium (reference) | 1 (reference) | 1 (reference) | 1 (reference) | 1 (reference) | 1 (reference) | 1 (reference) | 1 (reference) | 1 (reference) |
| Medium low HR, 95% CI | 1.31 (0.93, 1.85) | 1.21 (0.78, 1.88) | 1.20 (0.85, 1.69) | 1.16 (0.75, 1.80) | 1.26 (0.88, 1.79) | 1.16 (0.73, 1.83) | 1.25 (0.88, 1.78) | 1.21 (0.77, 1.91) |
| Low HR, 95% CI | **3.04 (2.17, 4.26)** | **2.64 (1.71, 4.07)** | 2.69 (1.91, 3.77) | **2.51 (1.63, 3.87)** | **2.79 (1.96, 3.97)** | **2.46 (1.57, 3.86)** | **3.01 (2.13, 4.24)** | **2.64 (1.70, 4.10)** |
| **Demographic characteristics** |  |  |  |  |  |  |  |  |
| Age HR, 95% CI | 1.01 (0.99, 1.03) | 1.00 (0.98, 1.02) | 1.02 (1.01, 1.04) | 1.01 (0.98, 1.03) | 1.00 (0.98, 1.02) | 0.99 (0.97, 1.01) | 0.99 (0.97, 1.00) | 0.98 (0.95, 1.00) |
| Female sex HR, 95% CI | 1.12 (0.88, 1.42) | **1.38 (1.02, 1.86)** | 1.13 (0.89, 1.44) | 1.34 (0.99, 1.82) | 1.12 (0.88, 1.44) | 1.31 (0.95, 1.81) | 1.02 (0.80, 1.30) | 1.22 (0.89, 1.68) |
| Education HR, 95% CI | 1.04 (1.00, 1.08) | 1.00 (0.94, 1.05) | 1.04 (1.00, 1.08) | 1.00 (0.94, 1.05) | 1.03 (0.99, 1.08) | 0.99 (0.93, 1.04) | 1.03 (0.99, 1.07) | 0.99 (0.94, 1.05) |
| **Additional covariates** |  |  |  |  |  |  |  |  |
| ≥1 *APOE* ε4 allele HR, 95% CI |  |  | **2.24 (1.76, 2.87)** | **1.67 (1.21, 2.29)** |  |  |  |  |
| Cortical GM HR, 95% CI |  |  |  |  | **0.62 (0.49, 0.80)** | **0.72 (0.53, 0.98)** |  |  |
| Hippocampal vol HR, 95% CI |  |  |  |  |  |  | **0.35 (0.27, 0.46)** | **0.40 (0.27, 0.59)** |

Note: Abbreviations: CI = Confidence Interval, HR = Hazard Ratio

### Supplemental Table S11. Cox model results for executive functioning at baseline and 6-month change (as in Table 2, left columns) plus each of Entorhinal thickness, Temporal lobe thickness, and Medial temporal lobe thickness

|  | **Executive Functioning** | | **Entorhinal cortex thickness** | | **Temporal lobe thickness** | | **Medial Temporal Lobe Thickness** | |
| --- | --- | --- | --- | --- | --- | --- | --- | --- |
|  | **All**  **(n = 764)** | **Amyloid + (n=408)** | **All**  **(n = 764)** | **Amyloid + (n=408)** | **All**  **(n = 764)** | **Amyloid + (n=408)** | **All**  **(n = 764)** | **Amyloid + (n=408)** |
| **6 month change categories** |  |  |  |  |  |  |  |  |
| Improved a lot HR, 95% CI | **0.49 (0.31, 0.79)** | **0.57 (0.33, 0.98)** | **0.45 (0.28, 0.73)** | 0.62 (0.35, 1.09) | **0.49 (0.31, 0.80)** | 0.65 (0.39, 1.15) | **0.49 (0.30, 0.78)** | 0.65 (0.36, 1.14) |
| Improved a little HR, 95% CI | **0.74 (0.55, 1.00)** | 0.80 (0.54, 1.19) | **0.73 (0.54, 0.99)** | 0.81 (0.54, 1.22) | **0.71 (0.52, 0.96)** | 0.79 (0.53, 1.18) | **0.71 (0.52, 0.96)** | 0.81 (0.54, 1.21) |
| Minimal change (reference) | 1 (reference) | 1 (reference) | 1 (reference) | 1 (reference) | 1 (reference) | 1 (reference) | 1 (reference) | 1 (reference) |
| A little worse HR, 95% CI | 1.17 (0.87, 1.58) | **1.56 (1.07, 2.29)** | 1.09 (0.80, 1.48) | 1.46 (0.98, 2.17) | 0.99 (0.73, 1.36) | 1.35 (0.90, 2.01) | 1.02 (0.75, 1.39) | 1.39 (0.93, 2.07) |
| A lot worse HR, 95% CI | 1.39 (0.86, 2.26) | 1.76 (0.94, 3/29) | 1.31 (0.80, 2.12) | 1.80 (0.96, 3.36) | 1.31 (0.80, 2.14) | 1.72 (0.92, 3.20) | 1.33 (0.82, 2.17) | 1.74 (0.93, 3.26) |
| **Baseline values** |  |  |  |  |  |  |  |  |
| High HR, 95% CI | **0.37 (0.24, 0.57)** | **0.37 (0.21, 0.66)** | **0.37 (0.23, 0.58)** | **0.40 (0.22, 0.74)** | **0.39 (0.25, 0.61)** | **0.40 (0.22, 0.74)** | **0.38 (0.24, 0.59)** | **0.39 (0.21, 0.73)** |
| Medium high HR, 95% CI | 0.69 (0.47, 1.01) | 0.71 (0.44, 1.15) | 0.69 (0.47, 1.03) | 0.88 (0.53, 1.46) | 0.70 (0.47, 1.05) | 0.85 (0.52, 1.40) | 0.68 (0.46, 1.01) | 0.84 (0.51, 1.39) |
| Medium (reference) | 1 (reference) | 1 (reference) | 1 (reference) | 1 (reference) | 1 (reference) | 1 (reference) | 1 (reference) | 1 (reference) |
| Medium low HR, 95% CI | 1.31 (0.93, 1.85) | 1.21 (0.78, 1.88) | 1.18 (0.83, 1.69) | 1.12 (0.71, 1.77) | 1.27 (0.89, 1.81) | 1.15 (0.73, 1.81) | 1.26 (0.88, 1.79) | 1.17 (0.74, 1.84) |
| Low HR, 95% CI | **3.04 (2.17, 4.26)** | **2.64 (1.71, 4.07)** | **2.79 (1.97, 3.95)** | **2.54 (1.63, 3.94)** | **2.51 (1.76, 3.58)** | **2.12 (1.34, 3.35)** | **2.62 (1.84, 3.72)** | **2.28 (1.45, 3.59)** |
| **Demographic characteristics** |  |  |  |  |  |  |  |  |
| Age HR, 95% CI | 1.01 (0.99, 1.03) | 1.00 (0.98, 1.02) | 1.00 (0.98, 1.01) | 0.99 (0.96, 1.01) | 0.99 (0.97, 1.01) | 0.98 (0.96, 1.01) | 1.00 (0.98, 1.01) | 0.99 (0.96, 1.01) |
| Female sex HR, 95% CI | 1.12 (0.88, 1.42) | **1.38 (1.02, 1.86)** | 1.12 (0.88, 1.43) | **1.37 (1.00, 1.87)** | 1.20 (0.94, 1.53) | **1.40 (1.03, 1.92)** | 1.19 (0.93, 1.51) | 1.43 (1.05, 1.95) |
| Education HR, 95% CI | 1.04 (1.00, 1.08) | 1.00 (0.94, 1.05) | 1.03 (0.98, 1.07) | 0.99 (0.94, 1.05) | 1.02 (0.98, 1.06) | 0.98 (0.93, 1.04) | 1.02 (0.98, 1.07) | 0.99 (0.93, 1.04) |
| **Additional covariates** |  |  | **0.48 (0.38, 0.60)** | **0.51 (0.37, 0.70)** |  |  |  |  |
| ER thickness HR, 95% CI |  |  | **<0.001** | **<0.001** |  |  |  |  |
| Temp. thickness HR, 95% CI |  |  |  |  | **<0.001** | **<0.001** |  |  |
| M temp thickness HR, 95% CI |  |  |  |  |  |  | **0.51 (0.40, 0.65)** | **0.56 (0.40, 0.77)** |

Note: Abbreviations: CI = Confidence Interval, HR = Hazard Ratio

### Supplemental Table S12. Cox model results for executive functioning at baseline and 6-month change (as in Table 2, left columns) plus each of Lateral Temporal Lobe Thickness, Frontal Lobe Thickness, and Cingulate Thickness

|  | **Executive Functioning** | | **Lateral Temporal Lobe Thickness** | | **Frontal lobe thickness** | | **Cingulate thickness** | |
| --- | --- | --- | --- | --- | --- | --- | --- | --- |
|  | **All**  **(n = 764)** | **Amyloid + (n=408)** | **All**  **(n = 764)** | **Amyloid + (n=408)** | **All**  **(n = 764)** | **Amyloid + (n=408)** | **All**  **(n = 764)** | **Amyloid + (n=408)** |
| **6 month change categories** |  |  |  |  |  |  |  |  |
| Improved a lot HR, 95% CI | **0.49 (0.31, 0.79)** | **0.57 (0.33, 0.98)** | **0.49 (0.30, 0.79)** | 0.64 (0.36, 1.13) | **0.46 (0.29, 0.74)** | 0.57 (0.32, 1.00) | **0.49 (0.30, 0.79)** | 0.61 (0.34, 1.07) |
| Improved a little HR, 95% CI | **0.74 (0.55, 1.00)** | 0.80 (0.54, 1.19) | **0.71 (0.52, 0.96)** | 0.78 (0.52, 1.17) | **0.69 (0.51, 0.94)** | 0.78 (0.52, 1.18) | **0.71 (0.52, 0.96)** | 0.84 (0.56, 1.26) |
| Minimal change (reference) | 1 (reference) | 1 (reference) | 1 (reference) | 1 (reference) | 1 (reference) | 1 (reference) | 1 (reference) | 1 (reference) |
| A little worse HR, 95% CI | 1.17 (0.87, 1.58) | **1.56 (1.07, 2.29)** | 0.99 (0.73, 1.36) | 1.35 (0.91, 2.02) | 1.01 (0.74, 1.38) | 1.42 (0.95, 2.12) | 1.06 (0.78, 1.44) | 1.49 (1.00, 2.21) |
| A lot worse HR, 95% CI | 1.39 (0.86, 2.26) | 1.76 (0.94, 3/29) | 1.32 (0.81, 2.16) | 1.74 (0.94, 3.25) | 1.39 (0.86, 2.27) | 1.75 (0.94, 3.29) | 1.39 (0.86, 2.27) | **1.91 (1.02, 3.57)** |
| **Baseline values** |  |  |  |  |  |  |  |  |
| High HR, 95% CI | **0.37 (0.24, 0.57)** | **0.37 (0.21, 0.66)** | **0.39 (0.25, 0.62)** | **0.39 (0.21, 0.73)** | **0.36 (0.23, 0.57)** | **0.36 (0.19, 0.66)** | **0.36 (0.23, 0.57)** | **0.32 (0.17, 0.60)** |
| Medium high HR, 95% CI | 0.69 (0.47, 1.01) | 0.71 (0.44, 1.15) | 0.72 (0.48, 1.06) | 0.83 (0.51, 1.36) | 0.71 (0.48. 1.05) | 0.79 (0.48, 1.30) | 0.69 (0.47, 1.03) | 0.74 (0.45, 1.21) |
| Medium (reference) | 1 (reference) | 1 (reference) | 1 (reference) | 1 (reference) | 1 (reference) | 1 (reference) | 1 (reference) | 1 (reference) |
| Medium low HR, 95% CI | 1.31 (0.93, 1.85) | 1.21 (0.78, 1.88) | 1.27 (0.89, 1.81) | 1.14 (0.73, 1.80) | 1.30 (0.91, 1.85) | 1.20 (0.76, 1.89) | 1.32 (0.93, 1.89) | 1.19 (0.76, 1.88) |
| Low HR, 95% CI | **3.04 (2.17, 4.26)** | **2.64 (1.71, 4.07)** | **2.56 (1.80, 3.66)** | **2.12 (1.34, 3.35)** | **2.95 (2.08, 4.19)** | **2.52 (1.62, 3.94)** | **3.08 (2.15, 4.32)** | **2.53 (1.62, 3.96)** |
| **Demographic characteristics** |  |  |  |  |  |  |  |  |
| Age HR, 95% CI | 1.01 (0.99, 1.03) | 1.00 (0.98, 1.02) | 0.99 (0.97, 1.01) | 0.98 (0.96, 1.00) | 1.00 (0.98, 1.02) | 0.99 (0.97, 1.01) | 1.01 (0.99, 1.03) | 1.00 (0.97, 1.02) |
| Female sex HR, 95% CI | 1.12 (0.88, 1.42) | **1.38 (1.02, 1.86)** | 1.20 (0.94, 1.54) | 1.38 (1.01, 1.89) | 1.27 (0.99, 1.62) | **1.48 (1.08, 2.03)** | 1.22 (0.95, 1.55) | **1.44 (1.06, 1.96)** |
| Education HR, 95% CI | 1.04 (1.00, 1.08) | 1.00 (0.94, 1.05) | 1.02 (0.98, 1.06) | 0.98 (0.92, 1.03) | 1.04 (0.99, 1.08) | 0.99 (0.94, 1.05) | 1.04 (1.00, 1.08) | 1.00 (0.94, 1.05) |
| **Additional covariates** |  |  |  |  |  |  |  |  |
| L Temp thickness HR, 95% CI |  |  | **0.46 (0.35, 0.59)** | **0.46 (0.33, 0.64)** |  |  |  |  |
| Frontal thickness HR, 95% CI |  |  |  |  | **0.68 (0.53, 0.88)** | **0.69 (0.50, 0.93)** |  |  |
| Cing. thickness HR, 95% CI |  |  |  |  |  |  | **0.73 (0.58, 0.93)** | **0.71 (0.53, 0.97)** |

Note: Abbreviations: CI = Confidence Interval, HR = Hazard Ratio

### Supplemental Table S13. Cox model results for executive functioning at baseline and 6-month change (as in Table 2, left columns) plus each of Parietal Lobe Thickness, Occipital Lobe Thickness, and Sensory / Motor Cortical Thickness

|  | **Executive Functioning** | | **Parietal Lobe Thickness** | | **Occipital lobe thickness** | | **Sensory Motor Cortical Thickness** | |
| --- | --- | --- | --- | --- | --- | --- | --- | --- |
|  | **All**  **(n = 764)** | **Amyloid + (n=408)** | **All**  **(n = 764)** | **Amyloid + (n=408)** | **All**  **(n = 764)** | **Amyloid + (n=408)** | **All**  **(n = 764)** | **Amyloid + (n=408)** |
| **6 month change categories** |  |  |  |  |  |  |  |  |
| Improved a lot HR, 95% CI | **0.49 (0.31, 0.79)** | **0.57 (0.33, 0.98)** | **0.51 (0.32, 0.83)** | 0.63 (0.36, 1.12) | **0.49 (0.30, 0.79)** | 0.62 (0.35, 1.10) | **0.47 (0.29, 0.77)** | 0.60 (0.34, 1.05) |
| Improved a little HR, 95% CI | **0.74 (0.55, 1.00)** | 0.80 (0.54, 1.19) | **0.73 (0.54, 0.99)** | 0.80 (0.53, 1.20) | **0.70 (0.52, 0.95)** | 0.79 (0.53, 1.19) | **0.71 (0.52, 0.96)** | 0.81 (0.54, 1.22) |
| Minimal change (reference) | 1 (reference) | 1 (reference) | 1 (reference) | 1 (reference) | 1 (reference) | 1 (reference) | 1 (reference) | 1 (reference) |
| A little worse HR, 95% CI | 1.17 (0.87, 1.58) | **1.56 (1.07, 2.29)** | 0.99 (0.73, 1.36) | 1.39 (0.93, 2.08) | 1.04 (0.76, 1.42) | 1.45 (0.97, 2.17) | 1.06 (0.78, 1.45) | **1.49 (1.00, 2.23)** |
| A lot worse HR, 95% CI | 1.39 (0.86, 2.26) | 1.76 (0.94, 3/29) | 1.38 (0.85, 2.25) | 1.87 (1.00, 3.50) | 1.41 (0.87, 2.30) | 1.89 (1.01, 3.53) | 1.43 (0.88, 2.33) | 1.87 (1.00, 3.50) |
| **Baseline values** |  |  |  |  |  |  |  |  |
| High HR, 95% CI | **0.37 (0.24, 0.57)** | **0.37 (0.21, 0.66)** | **0.39 (0.25, 0.62)** | **0.40 (0.21, 0.74)** | **0.37 (0.24, 0.59)** | **0.37 (0.20, 0.69)** | **0.36 (0.23, 0.57)** | **0.36 (0.19, 0.66)** |
| Medium high HR, 95% CI | 0.69 (0.47, 1.01) | 0.71 (0.44, 1.15) | 0.73 (0.49, 1.08) | 0.85 (0.52, 1.39) | 0.70 (0.47, 1.04) | 0.78 (0.47, 1.27) | 0.70 (0.47, 1.04) | 0.76 (0.46, 1.24) |
| Medium (reference) | 1 (reference) | 1 (reference) | 1 (reference) | 1 (reference) | 1 (reference) | 1 (reference) | 1 (reference) | 1 (reference) |
| Medium low HR, 95% CI | 1.31 (0.93, 1.85) | 1.21 (0.78, 1.88) | 1.32 (0.93, 1.89) | 1.24 (0.79, 1.95) | 1.32 (0.93, 1.88) | 1.23 (0.78, 1.93) | 1.32 (0.92, 1.88) | 1.22 (0.77, 1.91) |
| Low HR, 95% CI | **3.04 (2.17, 4.26)** | **2.64 (1.71, 4.07)** | **2.78 (1.96, 3.94)** | **2.37 (1.52, 3.69)** | **3.05 (2.15, 4.33)** | **2.56 (1.63, 4.00)** | **3.12 (2.20, 4.42)** | **2.66 (1.71, 4.14)** |
| **Demographic characteristics** |  |  |  |  |  |  |  |  |
| Age HR, 95% CI | 1.01 (0.99, 1.03) | 1.00 (0.98, 1.02) | 1.00 (0.98, 1.02) | 0.99 (0.97, 1.01) | 1.01 (0.99, 1.02) | 0.99 (0.97, 1.02) | 1.00 (0.99, 1.02) | 0.99 (0.97, 1.02) |
| Female sex HR, 95% CI | 1.12 (0.88, 1.42) | **1.38 (1.02, 1.86)** | 1.24 (0.97, 1.58) | **1.39 (1.02, 1.90)** | 1.21 (0.95, 1.55) | **1.41 (1.03, 1.93)** | 1.24 (0.97, 1.59) | **1.43 (1.05, 1.95)** |
| Education HR, 95% CI | 1.04 (1.00, 1.08) | 1.00 (0.94, 1.05) | 1.03 (0.99, 1.07) | 0.98 (0.92, 1.03) | 1.03 (0.99, 1.08) | 0.99 (0.93, 1.04) | 1.04 (1.00, 1.08) | 0.99 (0.94, 1.05) |
| **Additional covariates** |  |  |  |  |  |  |  |  |
| Parietal thickness HR, 95% CI |  |  | **0.52 (0.41, 0.67)** | **0.49 (0.36, 0.68)** |  |  |  |  |
| Occipit. thickness HR, 95% CI |  |  |  |  | 0.81 (0.64, 1.03) | 0.79 (0.59, 1.07) |  |  |
| Sen/Mot thickness HR, 95% CI |  |  |  |  |  |  | 0.83 (0.64, 1.07) | 0.85 (0.62, 1.16) |

Note: Abbreviations: CI = Confidence Interval, HR = Hazard Ratio

### Supplemental Table S14. Cox model results for language at baseline and 6-month change (as in Table 2, left columns) plus each of *APOE* genotype, Cortical Gray Matter Volume, and Hippocampal Volume

|  | **Language** | | ***APOE* genotype** | | **Cortical thickness** | | **Hippocampal volume** | |
| --- | --- | --- | --- | --- | --- | --- | --- | --- |
|  | **All**  **(n = 764)** | **Amyloid + (n=408)** | **All**  **(n = 764)** | **Amyloid + (n=408)** | **All**  **(n = 764)** | **Amyloid + (n=408)** | **All**  **(n = 764)** | **Amyloid + (n=408)** |
| **6 month change categories** |  |  |  |  |  |  |  |  |
| Improved a lot HR, 95% CI | **0.50 (0.29, 0.87)** | **0.45 (0.23, 0.89)** | **0.53 (0.31, 0.93)** | **0.47 (0.24, 0.93)** | **0.50 (0.29, 0.88)** | **0.45 (0.23, 0.89)** | 0.65 (0.38, 1.13) | 0.56 (0.28, 1.09) |
| Improved a little HR, 95% CI | **0.58 (0.42, 0.81)** | **0.53 (0.35, 0.80)** | **0.55 (0.39, 0.76)** | **0.50 (0.33, 0.76)** | **0.56 (0.40, 0.78)** | **0.48 (0.31, 0.75)** | **0.56 (0.40, 0.79)** | **0.46 (0.30, 0.72)** |
| Minimal change (reference) | 1 (reference) | 1 (reference) | 1 (reference) | 1 (reference) | 1 (reference) | 1 (reference) | 1 (reference) | 1 (reference) |
| A little worse HR, 95% CI | 1.33 (0.99, 1.77) | 1.05 (0.73, 1.52) | 1.21 (0.91, 1.62) | 0.99 (0.69, 1.43) | 1.21 (0.90, 1.63) | 0.99 (0.68, 1.44) | 1.24 (0.93, 1.67) | 0.98 (0.68, 1.42) |
| A lot worse HR, 95% CI | **2.28 (1.51, 3.46)** | **2.02 (1.19, 3.43)** | **2.16 (1.43, 3.27)** | **1.85 (1.09, 3.15)** | **2.11 (1.38, 3.22)** | **1.82 (1.06, 3.12)** | **2.07 (1.35, 3.18)** | **1.82 (1.06, 3.12)** |
| **Baseline values** |  |  |  |  |  |  |  |  |
| High HR, 95% CI | **0.26 (0.16, 0.41)** | **0.28 (0.16, 0.51)** | **0.27 (0.17, 0.43)** | **0.28 (0.16, 0.51)** | **0.25 (0.16, 0.41)** | **0.24 (0.13, 0.45)** | **0.28 (0.17, 0.45)** | **0.25 (0.13, 0.47)** |
| Medium high HR, 95% CI | 0.71 (0.49, 1.02) | 0.68 (0.44, 1.05) | **0.67 (0.47, 0.97)** | 0.67 (0.43, 1.04) | 0.70 (0.49, 1.02) | 0.66 (0.42, 1.04) | 0.75 (0.52, 1.08) | **0.63 (0.41, 0.99)** |
| Medium (reference) | 1 (reference) | 1 (reference) | 1 (reference) | 1 (reference) | 1 (reference) | 1 (reference) | 1 (reference) | 1 (reference) |
| Medium low HR, 95% CI | 1.00 (0.70, 1.43) | 0.95 (0.60, 1.50) | 1.09 (0.76, 1.56) | 1.02 (0.65, 1.61) | 1.07 (0.74, 1.55) | 0.89 (0.56, 1.43) | 0.97 (0.67, 1.41) | 0.81 (0.50, 1.31) |
| Low HR, 95% CI | **2.21 (1.56, 3.12)** | **2.22 (1.44, 3.42)** | **1.97 (1.39, 2.79)** | **2.21 (1.44, 3.40)** | **2.04 (1.44, 2.88)** | **1.91 (1.23, 2.94)** | **1.99 (1.40, 2.82)** | **1.76 (1.13, 2.73)** |
| **Demographic characteristics** |  |  |  |  |  |  |  |  |
| Age HR, 95% CI | 1.01 (1.00, 1.03) | 1.00 (0.98, 1.02) | **1.02 (1.01, 1.04)** | 1.01 (0.99, 1.03) | 1.00 (0.98, 1.02) | 0.99 (0.96, 1.01) | 0.99 (0.98, 1.01) | 0.98 (0.96, 1.00) |
| Female sex HR, 95% CI | 1.22 (0.96, 1.55) | 1.28 (0.95, 1.73) | 1.25 (0.98, 1.59) | 1.27 (0.94, 1.72) | 1.14 (0.89, 1.46) | 1.16 (0.84, 1.58) | 1.10 (0.86, 1.41) | 1.15 (0.85, 1.58) |
| Education HR, 95% CI | **1.05 (1.01, 1.09)** | 1.02 (0.97, 1.08) | 1.05 (1l.01, 1.09) | 1.02 (0.97, 1.08) | 1.04 (1.00, 1.09) | 1.01 (0.96, 1.07) | 1.03 (0.99, 1.08) | 1.01 (0.95, 1.07) |
| **Additional covariates** |  |  |  |  |  |  |  |  |
| ≥1 *APOE* ε4 allele HR, 95% CI |  |  | **2.32 (1.81, 2.96)** | **1.64 (1.19, 2.25)** |  |  |  |  |
| Cortical GM HR, 95% CI |  |  |  |  | **0.53 (0.42, 0.69)** | **0.56 (0.42, 0.76)** |  |  |
| Hippocampal vol HR, 95% CI |  |  |  |  |  |  | **0.40 (0.30, 0.51)** | **0.40 (0.28, 0.58)** |

Note: Abbreviations: CI = Confidence Interval, HR = Hazard Ratio

### Supplemental Table S15. Cox model results for language at baseline and 6-month change (as in Table 2, left columns) plus each of Entorhinal thickness, Temporal lobe thickness, and Medial temporal lobe thickness

|  | **Language** | | **Entorhinal thickness** | | **Temporal thickness** | | **Medial Temporal Lobe Thickness** | |
| --- | --- | --- | --- | --- | --- | --- | --- | --- |
|  | **All**  **(n = 764)** | **Amyloid + (n=408)** | **All**  **(n = 764)** | **Amyloid + (n=408)** | **All**  **(n = 764)** | **Amyloid + (n=408)** | **All**  **(n = 764)** | **Amyloid + (n=408)** |
| **6 month change categories** |  |  |  |  |  |  |  |  |
| Improved a lot HR, 95% CI | **0.50 (0.29, 0.87)** | **0.45 (0.23, 0.89)** | 0.60 (0.34. 1.04) | 0.53 (0.27, 1.05) | 0.59 (0.34, 1.03) | 0.55 (0.28, 1.09) | 0.59 (0.34, 1.03) | 0.55 (0.28, 1.08) |
| Improved a little HR, 95% CI | **0.58 (0.42, 0.81)** | **0.53 (0.35, 0.80)** | **0.56 (0.40, 0.78)** | **0.50 (0.32, 0.78)** | **0.59 (0.42, 0.83)** | **0.53 (0.34, 0.82)** | **0.57 (0.41, 0.79)** | **0.50 (0.32, 0.77)** |
| Minimal change (reference) | 1 (reference) | 1 (reference) | 1 (reference) | 1 (reference) | 1 (reference) | 1 (reference) | 1 (reference) | 1 (reference) |
| A little worse HR, 95% CI | 1.33 (0.99, 1.77) | 1.05 (0.73, 1.52) | 1.24 (0.92, 1.66) | 1.05 (0.73, 1.53) | 1.22 (0.91, 1.64) | 1.01 (0.70, 1.47) | 1.21 (0.90, 1.63) | 1.01 (0.70, 1.47) |
| A lot worse HR, 95% CI | **2.28 (1.51, 3.46)** | **2.02 (1.19, 3.43)** | **2.09 (1.36, 3.21)** | **1.91 (1.11, 3.29)** | **2.05 (1.34, 3.13)** | **1.90 (1.11, 3.26)** | **2.10 (1.37, 3.21)** | **1.91 (1.11, 3.29)** |
| **Baseline values** |  |  |  |  |  |  |  |  |
| High HR, 95% CI | **0.26 (0.16, 0.41)** | **0.28 (0.16, 0.51)** | **0.27 (0.17, 0.44)** | **0.26 (0.14, 0.50)** | **0.26 (0.16, 0.41)** | **0.24 (0.13, 0.44)** | **0.26 (0.16, 0.42)** | **0.24 (0.13, 0.46)** |
| Medium high HR, 95% CI | 0.71 (0.49, 1.02) | 0.68 (0.44, 1.05) | 0.79 (0.54, 1.14) | 0.66 (0.43, 1.04) | 0.72 (0.50, 1.05) | **0.61 (0.39, 0.96)** | 0.73 (0.50, 1.06) | **0.60 (0.38, 0.94)** |
| Medium (reference) | 1 (reference) | 1 (reference) | 1 (reference) | 1 (reference) | 1 (reference) | 1 (reference) | 1 (reference) | 1 (reference) |
| Medium low HR, 95% CI | 1.00 (0.70, 1.43) | 0.95 (0.60, 1.50) | 0.96 (0.66, 1.39) | 0.82 (0.51, 1.32) | 0.93 (0.64, 1.35) | 0.76 (0.47, 1.23) | 0.96 (0.66, 1.39) | 0.78 (0.48, 1.26) |
| Low HR, 95% CI | **2.21 (1.56, 3.12)** | **2.22 (1.44, 3.42)** | **1.91 (1.34, 2.72)** | **1.64 (1.05, 2.58)** | **1.78 (1.25, 2.53)** | 1.41 (0.89, 2.23) | **1.85 (1.30, 2.63)** | 1.47 (0.93, 2.34) |
| **Demographic characteristics** |  |  |  |  |  |  |  |  |
| Age HR, 95% CI | 1.01 (1.00, 1.03) | 1.00 (0.98, 1.02) | 1.00 (0.99, 1.02) | 0.99 (0.97, 1.01) | 1.00 (0.98, 1.02) | 0.99 (0.96, 1.01) | 1.00 (0.99, 1.02) | 0.99 (0.97, 1.01) |
| Female sex HR, 95% CI | 1.22 (0.96, 1.55) | 1.28 (0.95, 1.73) | 1.22 (0.96, 1.56) | 1.33 (0.98, 1.81) | 1.26 (0.99, 1.61) | 1.36 (1.00, 1.85) | 1.27 (0.99, 1.61) | **1.39 (1.02, 1.89)** |
| Education HR, 95% CI | **1.05 (1.01, 1.09)** | 1.02 (0.97, 1.08) | 1.03 (0.99, 1.07) | 1.01 (0.95, 1.06) | 1.03 (0.98, 1.07) | 1.00 (0.94, 1.05) | 1.03 (0.99, 1.07) | 1.00 (0.95, 1.06) |
| **Additional covariates** |  |  |  |  |  |  |  |  |
| ER thickness HR, 95% CI |  |  | **0.52 (0.41, 0.65)** | **0.52 (0.38, 0.71)** |  |  |  |  |
| Temp. thickness HR, 95% CI |  |  |  |  | **0.45 (0.35, 0.57)** | **0.41 (0.29, 0.57)** |  |  |
| M Temp thickness HR, 95% CI |  |  |  |  |  |  | **0.52 (0.41, 0.65)** | **0.48 (0.35, 0.66)** |

Note: Abbreviations: CI = Confidence Interval, HR = Hazard Ratio

### Supplemental Table S16. Cox model results for language at baseline and 6-month change (as in Table 2, left columns) plus each of Lateral Temporal Lobe Thickness, Frontal Lobe Thickness, and Cingulate Thickness

|  | **Language** | | **Lateral temporal lobe thickness** | | **Frontal lobe thickness** | | **Cingulate thickness** | |
| --- | --- | --- | --- | --- | --- | --- | --- | --- |
|  | **All**  **(n = 764)** | **Amyloid + (n=408)** | **All**  **(n = 764)** | **Amyloid + (n=408)** | **All**  **(n = 764)** | **Amyloid + (n=408)** | **All**  **(n = 764)** | **Amyloid + (n=408)** |
| **6 month change categories** |  |  |  |  |  |  |  |  |
| Improved a lot HR, 95% CI | **0.50 (0.29, 0.87)** | **0.45 (0.23, 0.89)** | **0.58 (0.33, 1.00)** | 0.53 (0.27, 1.05) | **0.53 (0.30, 0.91)** | **0.47 (0.24, 0.93)** | **0.50 (0.29, 0.88)** | **0.47 (0.24, 0.92)** |
| Improved a little HR, 95% CI | **0.58 (0.42, 0.81)** | **0.53 (0.35, 0.80)** | **0.60 (0.43, 0.85)** | **0.54 (0.35, 0.83)** | **0.53 (0.38, 0.75)** | **0.49 (0.31, 0.75)** | **0.56 (0.40, 0.78)** | **0.48 (0.31, 0.74)** |
| Minimal change (reference) | 1 (reference) | 1 (reference) | 1 (reference) | 1 (reference) | 1 (reference) | 1 (reference) | 1 (reference) | 1 (reference) |
| A little worse HR, 95% CI | 1.33 (0.99, 1.77) | 1.05 (0.73, 1.52) | 1.24 (0.92, 1.67) | 1.02 (0.70, 1.47) | 1.23 (0.91, 1.65) | 1.01 (0.70, 1.47) | 1.24 (0.92, 1.67) | 0.99 (0.68, 1.43) |
| A lot worse HR, 95% CI | **2.28 (1.51, 3.46)** | **2.02 (1.19, 3.43)** | **2.04 (1.34, 3.12)** | **1.87 (1.09, 3.21)** | **2.15 (1.41, 3.29)** | **1.81 (1.05, 3.10)** | **2.15 (1.41, 3.28)** | **1.77 (1.03, 3.04)** |
| **Baseline values** |  |  |  |  |  |  |  |  |
| High HR, 95% CI | **0.26 (0.16, 0.41)** | **0.28 (0.16, 0.51)** | **0.25 (0.16, 0.41)** | **0.24 (0.12, 0.44(** | **0.23 (0.14, 0.37)** | **0.23 (0.12, 0.42)** | **0.24 (0.15, 0.38)** | **0.23 (0.12, 0.43)** |
| Medium high HR, 95% CI | 0.71 (0.49, 1.02) | 0.68 (0.44, 1.05) | 0.71 (0.49, 1.03) | **0.62 (0.40, 0.98)** | **0.66 (0.46, 0.96)** | **0.62 (0.40, 0.97)** | 0.69 (0.48, 1.00) | 0.64 (0.41, 1.01) |
| Medium (reference) | 1 (reference) | 1 (reference) | 1 (reference) | 1 (reference) | 1 (reference) | 1 (reference) | 1 (reference) | 1 (reference) |
| Medium low HR, 95% CI | 1.00 (0.70, 1.43) | 0.95 (0.60, 1.50) | 0.92 (0.64, 1.34) | 0.77 (0.48, 1.25) | 1.05 (0.73, 1.52) | 0.85 (0.53, 1.37) | 1.05 (0.73, 1.52) | 0.86 (0.54, 1.39) |
| Low HR, 95% CI | **2.21 (1.56, 3.12)** | **2.22 (1.44, 3.42)** | **1.79 (1.26, 2.54)** | 1.47 (0.94, 2.31) | **2.06 (1.45, 2.93)** | **1.79 (1.14, 2.80)** | **2.15 (1.52, 3.05)** | **1.93 (1.24, 3.01)** |
| **Demographic characteristics** |  |  |  |  |  |  |  |  |
| Age HR, 95% CI | 1.01 (1.00, 1.03) | 1.00 (0.98, 1.02) | 1.00 (0.98, 1.01) | 0.98 (0.96, 1.01) | 1.00 (0.98, 1.02) | 0.99 (0.97, 1.02) | 1.01 (1.00, 1.03) | 1.00 (0.98, 1.03) |
| Female sex HR, 95% CI | 1.22 (0.96, 1.55) | 1.28 (0.95, 1.73) | 1.26 (0.99, 1.61) | 1.32 (0.97, 1.80) | **1.35 (1.05, 1.72)** | **1.40 (1.09, 1.90)** | **1.31 (1.02, 1.67)** | **1.39 (1.02, 1.89)** |
| Education HR, 95% CI | **1.05 (1.01, 1.09)** | 1.02 (0.97, 1.08) | 1.03 (0.98, 1.07) | 0.99 (0.94, 1.05) | **1.05 (1.01, 1.09)** | 1.02 (0.96, 1.08) | **1.05 (1.01, 1.10)** | 1.03 (0.97, 1.09) |
| **Additional covariates** |  |  |  |  |  |  |  |  |
| L temp thickness HR, 95% CI |  |  | **0.44 (0.34, 0.56)** | **041 (0.30, 0.57)** |  |  |  |  |
| Frontal thickness HR, 95% CI |  |  |  |  | **0.61 (0.48, 0.78)** | **0.61 (0.45, 0.82)** |  |  |
| Cing, thickness HR, 95% CI |  |  |  |  |  |  | **0.70 (0.56, 0.88)** | **0.66 (0.49, 0.90)** |

Note: Abbreviations: CI = Confidence Interval, HR = Hazard Ratio

### Supplemental Table S17. Cox model results for language at baseline and 6-month change (as in Table 2, left columns) plus each of Parietal Lobe Thickness, Occipital Lobe Thickness, and Sensory / Motor Cortical Thickness

|  | **Language** | | **Parietal Lobe Thickness** | | **Occipital lobe thickness** | | **Sensory Motor Cortical Thickness** | |
| --- | --- | --- | --- | --- | --- | --- | --- | --- |
|  | **All**  **(n = 764)** | **Amyloid + (n=408)** | **All**  **(n = 764)** | **Amyloid + (n=408)** | **All**  **(n = 764)** | **Amyloid + (n=408)** | **All**  **(n = 764)** | **Amyloid + (n=408)** |
| **6 month change categories** |  |  |  |  |  |  |  |  |
| Improved a lot HR, 95% CI | **0.50 (0.29, 0.87)** | **0.45 (0.23, 0.89)** | **0.48 (0.27, 0.83)** | **0.46 (0.23, 0.90)** | **0.53 (0.31, 0.92)** | 0.53 (0.27, 1.04) | **0.52 (0.30, 0.90)** | **0.49 (0.25, 0.96)** |
| Improved a little HR, 95% CI | **0.58 (0.42, 0.81)** | **0.53 (0.35, 0.80)** | **0.54 (0.39, 0.76)** | **0.48 (0.31, 0.75)** | **0.53 (0.38, 0.75)** | **0.47 (0.30, 0.72)** | **0.54 (0.39, 0.76)** | **0.50 (0.32, 0.77)** |
| Minimal change (reference) | 1 (reference) | 1 (reference) | 1 (reference) | 1 (reference) | 1 (reference) | 1 (reference) | 1 (reference) | 1 (reference) |
| A little worse HR, 95% CI | 1.33 (0.99, 1.77) | 1.05 (0.73, 1.52) | 1.19 (0.89, 1.60) | 0.99 (0.68, 1.43) | 1.23 (0.91, 1.65) | 1.02 (0.70, 1.48) | 1.26 (0.93, 1.69) | 1.02 (0.70, 1.48) |
| A lot worse HR, 95% CI | **2.28 (1.51, 3.46)** | **2.02 (1.19, 3.43)** | **2.02 (1.32, 3.09)** | **1.80 (1.05, 3.09)** | **2.25 (1.48, 3.45)** | **2.03 (1.19, 3.49)** | **2.22 (1.46, 3.40)** | **1.92 (1.12, 3.31)** |
| **Baseline values** |  |  |  |  |  |  |  |  |
| High HR, 95% CI | **0.26 (0.16, 0.41)** | **0.28 (0.16, 0.51)** | **0.24 (0.15, 0.39)** | **0.21 (0.11, 0.39)** | **0.23 (0.14, 0.38)** | **0.22 (0.12, 0.42)** | **0.24 (0.15, 0.38)** | **0.24 (0.13, 0.46)** |
| Medium high HR, 95% CI | 0.71 (0.49, 1.02) | 0.68 (0.44, 1.05) | **0.64 (0.44, 0.93)** | **0.54 (0.35, 0.85)** | **0.66 (0.46, 0.96)** | **0.59 (0.37, 0.92)** | **0.68 (0.47, 0.99)** | 0.65 (0.42, 1.02) |
| Medium (reference) | 1 (reference) | 1 (reference) | 1 (reference) | 1 (reference) | 1 (reference) | 1 (reference) | 1 (reference) | 1 (reference) |
| Medium low HR, 95% CI | 1.00 (0.70, 1.43) | 0.95 (0.60, 1.50) | 1.02 (0.71, 1.48) | 0.78 (0.48, 1.25) | 1.06 (0.74, 1.54) | 0.87 (0.54, 1.40) | 1.07 (0.74, 1.54) | 0.89 (0.56, 1.44) |
| Low HR, 95% CI | **2.21 (1.56, 3.12)** | **2.22 (1.44, 3.42)** | **1.97 (1.39, 2.80)** | **1.66 (1.06, 2.58)** | **2.10 (1.48, 2.97)** | **1.90 (1.22, 2.94)** | **2.16 (1.52, 3.06)** | **2.03 (1.31, 3.15)** |
| **Demographic characteristics** |  |  |  |  |  |  |  |  |
| Age HR, 95% CI | 1.01 (1.00, 1.03) | 1.00 (0.98, 1.02) | 1.00 (0.98, 1.02) | 0.99 (0.97, 1.01) | 1.00 (0.99, 1.02) | 0.99 (0.97, 1.01) | 1.01 (0.99, 1.02) | 1.0 (0.97, 1.02) |
| Female sex HR, 95% CI | 1.22 (0.96, 1.55) | 1.28 (0.95, 1.73) | **1.36 (1.06, 1.73)** | **1.38 (1.01, 1.87)** | **1.29 (1.01, 1.64)** | 1.34 (0.99, 1.82) | **1.32 (1.03, 1.69)** | 1.34 (0.99, 1.83) |
| Education HR, 95% CI | **1.05 (1.01, 1.09)** | 1.02 (0.97, 1.08) | 1.04 (1.00, 1.09) | 1.00 (0.95, 1.06) | 1.04 (1.00, 1.09) | 1.01 (0.95, 1.07) | 1.05 (1.01, 1.10) | 1.02 (0.96, 1.08) |
| **Additional covariates** |  |  |  |  |  |  |  |  |
| Parietal thickness HR, 95% CI |  |  | **0.46 (0.36, 0.58)** | **0.40 (0.29, 0.55)** |  |  |  |  |
| Occip. thickness HR, 95% CI |  |  |  |  | **0.65 (0.51, 0.83)** | **0.58 (0.43, 0.78)** |  |  |
| Sen/Mot thickness HR, 95% CI |  |  |  |  |  |  | 0.79 (0.62, 1.01) | 0.81 (0.60, 1.09) |

Note: Abbreviations: CI = Confidence Interval, HR = Hazard Ratio

### Supplemental Table S18. Cox model results for visuospatial at baseline and 6-month change (as in Table 2, left columns) plus each of *APOE* genotype, Cortical Gray Matter Volume, and Hippocampal Volume

|  | **Visuospatial alone** | | ***APOE* genotype** | | **Cortical gray volume** | | **Hippocampal volume** | |
| --- | --- | --- | --- | --- | --- | --- | --- | --- |
|  | **All**  **(n = 764)** | **Amyloid + (n=408)** | **All**  **(n = 764)** | **Amyloid + (n=408)** | **All**  **(n = 764)** | **Amyloid + (n=408)** | **All**  **(n = 764)** | **Amyloid + (n=408)** |
| **6 month change categories** |  |  |  |  |  |  |  |  |
| Improved a lot HR, 95% CI | 0.74 (0.39, 1.41) | 0.84 (0.39, 1.80) | 0.63 (0.33, 1.21) | 0.76 (0.35, 1.62) | 0.75 (0.40, 1.42) | 0.92 (0.43, 1.98) | 0.87 (0.45, 1.67) | 1.06 (0.49, 2.29) |
| Improved a little HR, 95% CI | **0.58 (0.42, 0.80)** | 0.67 (0.44, 1.02) | **0.56 (0.40, 0.77)** | **0.66 (0.43, 1.00)** | **0.58 (0.41, 0.80)** | **0.64 (0.41, 0.98)** | **0.64 (0.46, 0.88)** | **0.65 (0.42, 0.99)** |
| Minimal change (reference) | 1 (reference) | 1 (reference) | 1 (reference) | 1 (reference) | 1 (reference) | 1 (reference) | 1 (reference) | 1 (reference) |
| A little worse HR, 95% CI | 1.30 (0.95, 1.76) | 1.28 (0.88, 1.88) | 1.19 (0.87, 1.62) | 1.21 (0.82, 1.77) | 1.23 (0.90, 1.69) | 1.28 (0.86, 1.90) | 1.18 (0.86, 1.62) | 1.18 (0.80, 1.75) |
| A lot worse HR, 95% CI | **1.65 (1.03, 2.63)** | **1.92 (1.09, 3.37)** | 1.45 (0.91, 2.30) | **1.77 (1.01, 3.11)** | 1.46 (0.91, 2.34) | **1.85 (1.05, 3.26)** | 1.45 (0.91, 2.33) | 1.69 (0.96, 2.98) |
| **Baseline values** |  |  |  |  |  |  |  |  |
| High HR, 95% CI | **0.49 (0.37, 0.66)** | **0.57 (0.40, 0.83)** | **0.57 (0.42, 0.77)** | **0.60 (0.41, 0.87)** | **0.52 (0.38, 0.70)** | **0.57 (0.39, 0.84)** | **0.55 (0.40, 0.74)** | **0.58 (0.40, 0.85)** |
| Medium high HR, 95% CI | n/a | n/a | n/a | n/a | n/a | n/a | n/a | n/a |
| Medium (reference) | 1 (reference) | 1 (reference) | 1 (reference) | 1 (reference) | 1 (reference) | 1 (reference) | 1 (reference) | 1 (reference) |
| Medium low HR, 95% CI | 1.20 (0.84, 1.73) | 1.11 (0.68, 1.79) | 1.30 (0.91, 1.87) | 1.11 (0.69, 1.78) | 1.30 (0.90, 1.88) | 1.19 (0.73, 1.94) | 1.20 (0.83, 1.72) | 1.05 (0.65, 1.71) |
| Low HR, 95% CI | 1.33 (0.93, 1.90) | 1.43 (0.92, 2.23) | 1.41 (0.98, 2.02) | 1.52 (0.97, 2.38) | 1.18 (0.81, 1.70) | 1.22 (0.76, 1.94) | 1.24 (0.86, 1.79) | 1.21 (0.76, 1.93 |
| **Demographic characteristics** |  |  |  |  |  |  |  |  |
| Age HR, 95% CI | **1.03 (1.02, 1.05)** | 1.01 (0.99, 1.04) | **1.04 (1.03, 1.06)** | 1.02 (1.00, 1.05) | **1.02 (1.00, 1.04)** | 1.00 (0.98, 1.03) | 1.01 (0.44, 1.03) | 1.0 (0.97, 1.02) |
| Female sex HR, 95% CI | 1.05 (0.82, 1.33) | 1.23 (0.91, 1.66) | 1.10 (0.86, 1.40) | 1.22 (0.90, 1.65) | 0.99 (0.79, 1.27) | 1.11 (0.81, 1.51) | 0.95 (0.74. 1.22) | 1.09 (0.80, 1.49) |
| Education HR, 95% CI | 1.01 (0.97, 1.05) | 0.97 (0.92, 1.03) | 1.01 (0.97, 1.06) | 0.98 (0.93, 1.03) | 1.01 (0.96, 1.05) | 0.97 (0.92. 1.02) | 1.00 (0.96, 1.05) | 0.97 (0.92, 1.02) |
| **Additional covariates** |  |  |  |  |  |  |  |  |
| ≥1 *APOE* ε4 allele HR, 95% CI |  |  | **2.43 (1.91, 3.11)** | **1.60 (1.16, 2.20)** |  |  |  |  |
| Cortical GM HR, 95% CI |  |  |  |  | **0.50 (0.39, 0.64)** | **0.59 (0.43, 0.79)** |  |  |
| Hippocampal vol HR, 95% CI |  |  |  |  |  |  | **0.33 (0.25, 0.43)** | **0.38 (0.26, 0.55)** |

Note: Abbreviations: CI = Confidence Interval, HR = Hazard Ratio

### Supplemental Table S19. Cox model results for visuospatial at baseline and 6-month change (as in Table 2, left columns) plus each of Entorhinal thickness, Temporal lobe thickness, and Medial temporal lobe thickness

|  | **Visuospatial alone** | | **Entorhinal thickness** | | **Temporal thickness** | | **Medial temporal thickness** | |
| --- | --- | --- | --- | --- | --- | --- | --- | --- |
|  | **All**  **(n = 764)** | **Amyloid + (n=408)** | **All**  **(n = 764)** | **Amyloid + (n=408)** | **All**  **(n = 764)** | **Amyloid + (n=408)** | **All**  **(n = 764)** | **Amyloid + (n=408)** |
| **6 month change categories** |  |  |  |  |  |  |  |  |
| Improved a lot HR, 95% CI | 0.74 (0.39, 1.41) | 0.84 (0.39, 1.80) | 0.71 (0.37, 1.36) | 0.76 (0.36, 1.63) | 0.69 (0.36, 1.31) | 0.76 (0.36, 1.63) | 0.67 (0.35, 1.27) | 0.75 (0.35, 1.61) |
| Improved a little HR, 95% CI | **0.58 (0.42, 0.80)** | 0.67 (0.44, 1.02) | **0.65 (0.47, 0.90)** | 0.70 (0.45, 1.07) | **0.64 (0.46, 0.89)** | 0.75 (0.49, 1.16) | **0.65 (0.47, 0.90)** | 0.75 (0.48, 1.15) |
| Minimal change (reference) | 1 (reference) | 1 (reference) | 1 (reference) | 1 (reference) | 1 (reference) | 1 (reference) | 1 (reference) | 1 (reference) |
| A little worse HR, 95% CI | 1.30 (0.95, 1.76) | 1.28 (0.88, 1.88) | 1.24 (0.90, 1.70) | 1.19 (0.80, 1.77) | 1.21 (0.88, 1.66) | 1.18 (0.80, 1.76) | 1.22 (0.89, 1.67) | 1.17 (0.79, 1.74) |
| A lot worse HR, 95% CI | **1.65 (1.03, 2.63)** | **1.92 (1.09, 3.37)** | **1.67 (1.04, 2.68)** | 1.76 (1.00, 3.12) | 1.53 (0.96, 2.45) | 1.62 (0.92, 2.87) | **1.65 (1.03, 2.64)** | 1.72 (0.98, 3.05) |
| **Baseline values** |  |  |  |  |  |  |  |  |
| High HR, 95% CI | **0.49 (0.37, 0.66)** | **0.57 (0.40, 0.83)** | **0.55 (0.41, 0.75)** | **0.57 (0.39, 0.84)** | **0.52 (0.38, 0.70)** | **0.59 (0.40, 0.86)** | **0.54 (0.40, 0.73)** | **0.59 (0.41, 0.87)** |
| Medium high HR, 95% CI | n/a | n/a | n/a | n/a | n/a | n/a | n/a | n/a |
| Medium (reference) | 1 (reference) | 1 (reference) | 1 (reference) | 1 (reference) | 1 (reference) | 1 (reference) | 1 (reference) | 1 (reference) |
| Medium low HR, 95% CI | 1.20 (0.84, 1.73) | 1.11 (0.68, 1.79) | 1.32 (0.91, 1.91) | 1.27 (0.78, 2.07) | 1.22 (0.85, 1.76) | 1.12 (0.69, 1.83) | 1.24 (0.86, 1.79) | 1.16 (0.71, 1.89) |
| Low HR, 95% CI | 1.33 (0.93, 1.90) | 1.43 (0.92, 2.23) | 1.37 (0.94, 1.98) | 1.32 (0.84, 2.10) | 1.19 (0.83, 1.72) | 1.22 (0.77, 1.94) | 1.30 (0.90, 1.89) | 1.33 (0.84, 2.12) |
| **Demographic characteristics** |  |  |  |  |  |  |  |  |
| Age HR, 95% CI | **1.03 (1.02, 1.05)** | 1.01 (0.99, 1.04) | **1.02 (1.00, 1.04)** | 1.00 (0.98, 1.02) | 1.01 (0.99, 1.03) | 0.99 (0.97, 1.01) | 1.02 (1.00, 1.03) | 1.00 (0.98, 1.02) |
| Female sex HR, 95% CI | 1.05 (0.82, 1.33) | 1.23 (0.91, 1.66) | 1.05 (0.83, 1.35) | 1.23 (0.91, 1.67) | 1.13 (0.88, 1.44) | 1.29 (0.95, 1.74) | 1.12 (0.87, 1.43) | 1.31 (0.97, 1.78) |
| Education HR, 95% CI | 1.01 (0.97, 1.05) | 0.97 (0.92, 1.03) | 1.01 (0.96, 1.05) | 0.98 (0.93, 1.04) | 1.00 (0.96, 1.04) | 0.97 (0.92, 1.03) | 1.00 (0.96, 1.05) | 0.98 (0.92, 1.03) |
| **Additional covariates** |  |  |  |  |  |  |  |  |
| ER thickness HR, 95% CI |  |  | **0.44 (0.35, 0.55)** | **0.45 (0.33, 0.61)** |  |  |  |  |
| Temp. thickness HR, 95% CI |  |  |  |  | **0.36 (0.28, 0.46)** | **0.35 (0.25, 0.49)** |  |  |
| M temp. thickness HR, 95% CI |  |  |  |  |  |  | **0.43 (0.34, 0.55)** | **0.42 (0.31, 0.58)** |

Note: Abbreviations: CI = Confidence Interval, HR = Hazard Ratio

### Supplemental Table S20. Cox model results for visuospatial at baseline and 6-month change (as in Table 2, left columns) plus each of Lateral Temporal Lobe Thickness, Frontal Lobe Thickness, and Cingulate Thickness

|  | **Visuospatial alone** | | **Lateral temporal lobe thickness** | | **Frontal lobe thickness** | | **Cingulate thickness** | |
| --- | --- | --- | --- | --- | --- | --- | --- | --- |
|  | **All**  **(n = 764)** | **Amyloid + (n=408)** | **All**  **(n = 764)** | **Amyloid + (n=408)** | **All**  **(n = 764)** | **Amyloid + (n=408)** | **All**  **(n = 764)** | **Amyloid + (n=408)** |
| **6 month change categories** |  |  |  |  |  |  |  |  |
| Improved a lot HR, 95% CI | 0.74 (0.39, 1.41) | 0.84 (0.39, 1.80) | 0.73 (0.38, 1.39) | 0.79 (0.39, 1.68) | 0.77 (0.40, 1.46) | 0.88 (0.41, 1.88) | 0.74 (0.39, 1.41) | 0.87 (0.41, 1.87) |
| Improved a little HR, 95% CI | **0.58 (0.42, 0.80)** | 0.67 (0.44, 1.02) | **0.62 (0.45, 0.86)** | 0.72 (0.47, 1.12) | **0.59 (0.43, 0.82)** | 0.66 (0.43, 1.02) | **0.56 (0.40, 0.78)** | **0.60 (0.39, 0.93)** |
| Minimal change (reference) | 1 (reference) | 1 (reference) | 1 (reference) | 1 (reference) | 1 (reference) | 1 (reference) | 1 (reference) | 1 (reference) |
| A little worse HR, 95% CI | 1.30 (0.95, 1.76) | 1.28 (0.88, 1.88) | 1.21 (0.88, 1.66) | 1.21 (0.82, 1.80) | 1.22 (0.89, 1.67) | 1.23 (0.83, 1.82) | 1.30 (0.95, 1.78) | 1.33 (0.90, 1.97) |
| A lot worse HR, 95% CI | **1.65 (1.03, 2.63)** | **1.92 (1.09, 3.37)** | 1.46 (0.91, 2.34) | 1.58 (0.89, 2.80) | 1.47 (0.92, 2.36) | 1.75 (0.99, 3.09) | 1.50 (0.93, 2.40) | 1.74 (0.99, 3.08) |
| **Baseline values** |  |  |  |  |  |  |  |  |
| High HR, 95% CI | **0.49 (0.37, 0.66)** | **0.57 (0.40, 0.83)** | **0.50 (0.37, 0.68)** | **0.58 (0.40, 0.85)** | **0.51 (0.38, 0.69)** | **0.58 (0.39, 0.84)** | **0.49 (0.36, 0.67)** | **0.55 (0.37, 0.80)** |
| Medium high HR, 95% CI | n/a | n/a | n/a | n/a | n/a | n/a | n/a | n/a |
| Medium (reference) | 1 (reference) | 1 (reference) | 1 (reference) | 1 (reference) | 1 (reference) | 1 (reference) | 1 (reference) | 1 (reference) |
| Medium low HR, 95% CI | 1.20 (0.84, 1.73) | 1.11 (0.68, 1.79) | 1.21 (0.84, 1.74) | 1.11 (0.68, 1.81) | 1.30 (0.90, 1.88) | 1.18 (0.72, 1.92) | 1.29 (0.89, 1.87) | 1.20 (0.74, 1.96) |
| Low HR, 95% CI | 1.33 (0.93, 1.90) | 1.43 (0.92, 2.23) | 1.12 (0.78, 1.62) | 1.16 (0.73, 1.84) | 1.24 (0.86, 1.79) | 1.30 (0.82, 2.05) | 1.29 (0.89, 1.87) | 1.39 (0.88, 2.20) |
| **Demographic characteristics** |  |  |  |  |  |  |  |  |
| Age HR, 95% CI | **1.03 (1.02, 1.05)** | 1.01 (0.99, 1.04) | 1.01 (0.99, 1.03) | 0.99 (0.97, 1.01) | **1.02 (1.01, 1.04)** | 1.01 (0.98, 1.03) | **1.04 (1.02, 1.05)** | 1.02 (1.00, 1.04) |
| Female sex HR, 95% CI | 1.05 (0.82, 1.33) | 1.23 (0.91, 1.66) | 1.12 (0.88, 1.44) | 1.25 (0.92, 1.70) | 1.15 (0.90, 1.48) | 1.31 (0.96, 1.79) | 1.12 (0.88, 1.44) | 1.27 (0.93, 1.73) |
| Education HR, 95% CI | 1.01 (0.97, 1.05) | 0.97 (0.92, 1.03) | 1.00 (0.96, 1.04) | 0.96 (0.91, 1.02) | 1.01 (0.97, 1.05) | 0.97 (0.92, 1.03) | 1.01 (0.97, 1.06) | 0.98 (0.93, 1.04) |
| **Additional covariates** |  |  |  |  |  |  |  |  |
| L Temp thickness HR, 95% CI |  |  | **0.35 (0.27, 0.45)** | **0.35 (0.25, 0.49)** |  |  |  |  |
| Frontal thickness HR, 95% CI |  |  |  |  | **0.63 (0.48, 0.81)** | **0.63 (0.46, 0.86)** |  |  |
| Cing. Thickness HR, 95% CI |  |  |  |  |  |  | **0.66 (0.52, 0.83)** | **0.66 (0.49, 0.90)** |

Note: Abbreviations: CI = Confidence Interval, HR = Hazard Ratio

### Supplemental Table S21. Cox model results for visuospatial at baseline and 6-month change (as in Table 2, left columns) plus each of Parietal Lobe Thickness, Occipital Lobe Thickness, and Sensory / Motor Cortical Thickness

|  | **Visuospatial alone** | | **Parietal lobe thickness** | | **Occipital lobe thickness** | | **Sensory Motor Cortical Thickness** | |
| --- | --- | --- | --- | --- | --- | --- | --- | --- |
|  | **All**  **(n = 764)** | **Amyloid + (n=408)** | **All**  **(n = 764)** | **Amyloid + (n=408)** | **All**  **(n = 764)** | **Amyloid + (n=408)** | **All**  **(n = 764)** | **Amyloid + (n=408)** |
| **6 month change categories** |  |  |  |  |  |  |  |  |
| Improved a lot HR, 95% CI | 0.74 (0.39, 1.41) | 0.84 (0.39, 1.80) | 0.79 (0.42, 1.50) | 0.90 (0.42, 1.91) | 0.77 (0.41, 1.47) | 0.88 (0.41, 1.88) | 0.75 (0.39, 1.43) | 0.83 (0.39, 1.77) |
| Improved a little HR, 95% CI | **0.58 (0.42, 0.80)** | 0.67 (0.44, 1.02) | **0.61 (0.44, 0.86)** | 0.69 (0.45. 1.07) | **0.59 (0.43, 0.82)** | 0.68 (0.44, 1.05) | **0.58 (0.42, 0.81)** | **0.64 (0.41, 0.98)** |
| Minimal change (reference) | 1 (reference) | 1 (reference) | 1 (reference) | 1 (reference) | 1 (reference) | 1 (reference) | 1 (reference) | 1 (reference) |
| A little worse HR, 95% CI | 1.30 (0.95, 1.76) | 1.28 (0.88, 1.88) | 1.17 (0.85, 1.61) | 1.15 (0.77, 1.72) | 1.24 (0.90, 1.70) | 1.26 (0.85, 1.86) | 1.27 (0.92, 1.74) | 1.27 (0.86, 1.88) |
| A lot worse HR, 95% CI | **1.65 (1.03, 2.63)** | **1.92 (1.09, 3.37)** | 1.42 (0.89, 2.28) | 1.62 (0.92, 2.86) | 1.52 (0.95, 2.43) | 1.70 (0.96, 3.01) | 1.55 (0.97, 2.49) | **1.82 (1.03, 3.21)** |
| **Baseline values** |  |  |  |  |  |  |  |  |
| High HR, 95% CI | **0.49 (0.37, 0.66)** | **0.57 (0.40, 0.83)** | **0.52 (0.38, 0.70)** | **0.56 (0.38, 0.82)** | **0.52 (0.38, 0.70)** | **0.59 (0.40, 0.86)** | **0.50 (0.36, 0.68)** | **0.57 (0.39, 0.83)** |
| Medium high HR, 95% CI | n/a | n/a | n/a | n/a | n/a | n/a | n/a | n/a |
| Medium (reference) | 1 (reference) | 1 (reference) | 1 (reference) | 1 (reference) | 1 (reference) | 1 (reference) | 1 (reference) | 1 (reference) |
| Medium low HR, 95% CI | 1.20 (0.84, 1.73) | 1.11 (0.68, 1.79) | 1.25 (0.86, 1.80) | 1.05 (0.64, 1.73) | 1.30 (0.90, 1.87) | 1.22 (0.75, 1.99) | 1.28 (0.88, 1.85) | 1.21 (0.74, 1.97) |
| Low HR, 95% CI | 1.33 (0.93, 1.90) | 1.43 (0.92, 2.23) | 1.15 (0.79, 1.66) | 1.15 (0.73, 1.82) | 1.24 (0.85, 1.79) | 1.32 (0.83, 2.08) | 1.26 (0.87, 1.82) | 1.36 (0.86, 2.15) |
| **Demographic characteristics** |  |  |  |  |  |  |  |  |
| Age HR, 95% CI | **1.03 (1.02, 1.05)** | 1.01 (0.99, 1.04) | **1.02 (1.00, 1.04)** | 1.00 (0.98, 1.02) | **1.03 (1.01, 1.05)** | 1.01 (0.99, 1.03) | **1.03 (1.01, 1.05)** | 1.01 (0.98, 1.03) |
| Female sex HR, 95% CI | 1.05 (0.82, 1.33) | 1.23 (0.91, 1.66) | 1.16 (0.91, 1.49) | 1.28 (0.94, 1.73) | 1.10 (0.86, 1.41) | 1.24 (0.91, 1.68) | 1.13 (0.88, 1.44) | 1.26 (0.93, 1.72) |
| Education HR, 95% CI | 1.01 (0.97, 1.05) | 0.97 (0.92, 1.03) | 1.00 (0.96, 1.05) | 0.96 (0.91, 1.02) | 1.00 (0.96, 1.05) | 0.97 (0.92, 1.02) | 1.01 (0.97, 1.05) | 0.97 (0.92, 1.03) |
| **Additional covariates** |  |  |  |  |  |  |  |  |
| Parietal thickness HR, 95% CI |  |  | **0.43 (0.34, 0.56)** | **0.42 (0.30, 0.57)** |  |  |  |  |
| Occip. thickness HR, 95% CI |  |  |  |  | **0.69 (0.54, 0.88)** | **0.66 (0.49, 0.90)** |  |  |
| Sen/Mot thickness HR, 95% CI |  |  |  |  |  |  | 0.80 (0.62, 1.04) | 0.79 (0.58, 1.08) |

Note: Abbreviations: CI = Confidence Interval, HR = Hazard Ratio

### Supplemental Table S22. Cox model results for memory without and with the CSF biomarker signature

|  | **Memory** | | **Biomarker signature** | |
| --- | --- | --- | --- | --- |
|  | **All**  **(n = 764)** | **Amyloid + (n=408)** | **All**  **(n = 764)** | **Amyloid + (n=408)** |
| **6-month change categories** |  |  |  |  |
| Improved HR, 95% CI | 0.76 (0.43, 1.34) | 0.62 (0.33, 1.15) | 0.85 (0.48, 1.49) | 0.70 (0.36, 1.36) |
| Minimal change (reference) | 1 (reference) | 1 (reference) | 1 (reference) | 1 (reference) |
| Worse HR, 95% CI | 1.34 (0.85, 2.11) | 1.07 (0.66, 1.74) | 1.23 (0.78, 1.95) | 1.03 (0.63, 1.69) |
| **Baseline value categories** |  |  |  |  |
| High HR, 95% CI | **0.14 (0.03, 0.62)** | 0.25 (0.05, 1.13) | **0.15 (0.03, 0.69)** | 0.24 (0.05, 1.09) |
| Medium high HR, 95% CI | 0.64 (0.29, 1.38) | 1.26 (0.50, 3.20) | 0.84 (0.38, 1.86) | 1.32 (0.52, 3.37) |
| Medium (reference) | 1 (reference) | 1 (reference) | 1 (reference) | 1 (reference) |
| Medium low HR, 95% CI | **1.87 (1.00, 3.46)** | 1.97 (0.98, 3.97) | 1.65 (0.89, 3.09) | 1.90 (0.94, 3.85) |
| Low HR, 95% CI | **2.99 (1.62, 5.52)** | **3.35 (1.67, 6.70)** | **2.67 (1.44, 4.94)** | **3.24 (1.62, 6.48)** |
| **Demographic characteristics** |  |  |  |  |
| Age HR, 95% CI | 1.00 (0.97, 1.03) | 1.00 (0.97, 1.03) | 1.01 (0.98, 1.03) | 1.00 (0.97, 1.04) |
| Female sex HR, 95% CI | 1.53 (0.98, 2.40) | **1.67 (1.02, 2.75)** | 1.42 (0.90, 2.23) | 1.63 (0.99, 2.69) |
| Education HR, 95% CI | 1.00 (0.93, 1.07) | 0.99 (0.92, 1.07) | 1.00 (0.94, 1.08) | 0.99 (0.92, 1.07) |
| CSF biomarker HR, 95% CI |  |  | **2.46 (1.31, 4.62)** | 1.69 (0.60, 4.74) |

Note: Abbreviations: CI = Confidence Interval, HR = Hazard Ratio

### Supplemental Table S23. Cox model results for executive functioning without and with the CSF biomarker signature

|  | **Executive Functioning Alone** | | **Biomarker signature** | |
| --- | --- | --- | --- | --- |
|  | **All**  **(n = 764)** | **Amyloid + (n=408)** | **All**  **(n = 764)** | **Amyloid + (n=408)** |
| **6-month change categories** |  |  |  |  |
| Improved HR, 95% CI | 0.60 (0.35, 1.03) | 0.69 (0.37, 1.27) | 0.69 (0.40, 1.19) | 0.73 (0.39, 1.35) |
| Minimal change (reference) | 1 (reference) | 1 (reference) | 1 (reference) | 1 (reference) |
| Worse HR, 95% CI | 1.39 (0.88, 2.21) | **1.86 (1.11, 3.12)** | 1.53 (0.95, 2.44) | **1.84 (1.09, 3.10)** |
| **Baseline value categories** |  |  |  |  |
| High HR, 95% CI | **0.34 (0.14, 0.80)** | 0.80 (0.29, 2.20) | 0.48 (0.20, 1.17) | 0.70 (0.25, 1.94) |
| Medium high HR, 95% CI | **0.38 (0.20, 0.75)** | 0.52 (0.25, 1.09) | **0.38 (0.20, 0.75)** | **0.46 (0.22, 0.97)** |
| Medium (reference) | 1 (reference) | 1 (reference) | 1 (reference) | 1 (reference) |
| Medium low HR, 95% CI | 0.66 (0.36, 1.22) | 0.95 (0.48, 1.87) | 0.67 (0.36, 1.25) | 0.80 (0.40, 1.61) |
| Low HR, 95% CI | 1.16 (0.63, 2.14) | 1.55 (0.80, 3.00) | 1.02 (0.56, 1.88) | 1.41 (0.73, 2.71) |
| **Demographic characteristics** |  |  |  |  |
| Age HR, 95% CI | 1.00 (0.97, 1.03) | 1.00 (0.97, 1.03) | 1.01 (0.98, 1.04) | 1.00 (0.97, 1.04) |
| Female sex HR, 95% CI | 1.45 (0.93, 2.27) | 1.46 (0.88, 2.42) | 1.37 (0.98, 2.18) | 1.49 (0.90, 2.48) |
| Education HR, 95% CI | 0.97 (0.90, 1.04) | 0.97 (0.90, 1.05) | 0.97 (0.90, 1.04) | 0.97 (0.90, 1.05) |
| CSF biomarker HR, 95% CI |  |  | **3.02 (1.66, 5.52)** | 2.16 (0.82, 5.70) |

Note: Abbreviations: CI = Confidence Interval, HR = Hazard Ratio

### Supplemental Table S24. Cox model results for language without and with the CSF biomarker signature

|  | **Language Alone** | | **Biomarker signature** | |
| --- | --- | --- | --- | --- |
|  | **All**  **(n = 764)** | **Amyloid + (n=408)** | **All**  **(n = 764)** | **Amyloid + (n=408)** |
| **6-month change categories** |  |  |  |  |
| Improved HR, 95% CI | **0.51 (0.29, 0.88)** | **0.43 (0.24, 0.78)** | **0.42 (0.24, 0.75)** | **0.32 (0.17, 0.61)** |
| Minimal change (reference) | 1 (reference) | 1 (reference) | 1 (reference) | 1 (reference) |
| Worse HR, 95% CI | 1.54 (0.97, 2.44) | 1.09 (0.66, 1.79) | 1.18 (0.73, 1.89) | 0.86 (0.51, 1.45) |
| **Baseline value categories** |  |  |  |  |
| High HR, 95% CI | 0.56 (0.22, 1.45) | 0.62 (0.22, 1.76) | 0.53 (0.21, 1.34) | 0.62 (0.22, 1.77) |
| Medium high HR, 95% CI | 0.93 (0.48, 1.79) | 0.75 (0.37, 1.52) | 0.70 (0.36, 1.37) | 0.60 (0.29, 1.26) |
| Medium (reference) | 1 (reference) | 1 (reference) | 1 (reference) | 1 (reference) |
| Medium low HR, 95% CI | 0.94 (0.51, 1.75) | 0.78 (0.40, 1.54) | 0.68 (0.36, 1.28) | 0.66 (0.33, 1.32) |
| Low HR, 95% CI | **2.10 (1.14, 3,86)** | **2.10 (1.05, 4.18)** | **1.89 (1.03, 3.48)** | **2.20 (1.11, 4.36)** |
| **Demographic characteristics** |  |  |  |  |
| Age HR, 95% CI | 0.99 (0.96, 1.02) | 0.98 (0.95, 1.01) | 0.99 (0.96, 1.02) | 0.98 (0.95, 1.01) |
| Female sex HR, 95% CI | 1.27 (0.80, 2.02) | 1.47 (0.88, 2.46) | 1.17 (0.73, 1.88) | 1.40 (0.83, 2.34) |
| Education HR, 95% CI | 1.02 (0.95, 1.10) | 1.04 (0.96, 1.13) | 1.03 (0.96, 1.12) | 1.06 (0.97, 1.15) |
| CSF biomarker HR, 95% CI |  |  | **3.93 (2.14, 7.19)** | **3.63 (1.30, 10.1)** |

Note: Abbreviations: CI = Confidence Interval, HR = Hazard Ratio

### Supplemental Table S25. Cox model results for visuospatial without and with the CSF biomarker signature

|  | **Visuospatial Alone** | | **Biomarker signature** | |
| --- | --- | --- | --- | --- |
|  | **All**  **(n = 764)** | **Amyloid + (n=408)** | **All**  **(n = 764)** | **Amyloid + (n=408)** |
| **6-month change categories** |  |  |  |  |
| Improved HR, 95% CI | 0.61 (0.35, 1.07) | 0.87 (0.48, 1.58) | 0.67 (0.38, 1.18) | 0.87 (0.48, 1.57) |
| Minimal change (reference) | 1 (reference) | 1 (reference) | 1 (reference) | 1 (reference) |
| Worse HR, 95% CI | 1.12 (0.69, 1.81) | 1.22 (0.73, 2.05) | 1.15 (0.71, 1.86) | 1.28 (0.76, 2.16) |
| **Baseline value categories** |  |  |  |  |
| High HR, 95% CI | 0.70 (0.41, 1.19) | 0.69 (0.39, 1.22) | 0.81 (0.48, 1.38) | 0.70 (0.39, 1.24) |
| Medium high HR, 95% CI | n/a | n/a | n/a | n/a |
| Medium (reference) | 1 (reference) | 1 (reference) | 1 (reference) | 1 (reference) |
| Medium low HR, 95% CI | 1.31 (0.71, 2.41) | 0.89 (0.46, 1.72) | 1.18 (0.64, 2.19) | 0.87 (0.45, 1.68) |
| Low HR, 95% CI | 1.24 (0.70, 2.21) | 0.91 (0.49, 1.68) | 1.04 (0.58, 1.85) | 0.85 (0.45, 1.58) |
| **Demographic characteristics** |  |  |  |  |
| Age HR, 95% CI | 1.01 (0.98, 1.03) | 1.00 (0.97, 1.04) | 1.01 (0.98, 1.04) | 1.01 (0.97, 1.04) |
| Female sex HR, 95% CI | 1.36 (0.87, 2.12) | 1.62 (0.99. 2.65) | 1.32 (0.84, 2.08) | 1.60 (0.98, 2.63) |
| Education HR, 95% CI | 0.99 (0.92, 1.06) | 0.99 (0.92, 1.06) | 1.00 (0.93, 1.07) | 0.99 (0.92, 1.07) |
| CSF biomarker HR, 95% CI |  |  | **3.28 (1.84, 5.85)** | 2.06 (0.81, 5.21) |

Note: Abbreviations: CI = Confidence Interval, HR = Hazard Ratio
